# Machine learning augmentation reduces prediction error in collective forecasting: development and validation across prediction markets

**DOI:** 10.1101/2023.01.19.23284578

**Authors:** Alexander Gruen, Karl R Mattingly, Ellen Morwitch, Frederik Bossaerts, Manning Clifford, Chad Nash, John P A Ioannidis, Anne-Louise Ponsonby

## Abstract

The recent COVID-19 crisis highlighted the inadequacy of human forecasting. We aim to leverage human prediction markets with real-time machine weighting of likely higher accuracy trades to improve performance. The crowd sourced Almanis prediction market longitudinal platform (n=1822) and Next Generation Social Science (NGS2) platform (n=103) were utilised. A 43-feature model predicted top quintile relative Brier accuracy scores in two out-of-sample datasets (p_*both*_<1×10^−9^). Trades graded as high machine accuracy quality vs. other trades had a greater AUC temporal gain from before to after trade. Hybrid human-machine forecasts had higher accuracy than human forecasts alone, particularly when the two systems disagreed by 5% or more for binary event prediction: the hybrid system demonstrating substantial AUC gains of 13.2%, p=1.35×10^−14^ and 13.8%, p=0.003 in the out-of-sample Almanis B and NGS2 datasets respectively. When discordant, the hybrid model was correct for COVID-19 event occurrence 72.7% of the time vs 27.3% for human-only models, p=0.007. This net classification benefit was replicated in the separate Almanis B dataset, p=2.4×10^−7^. Real-time machine classification followed by weighting human trades according to likely accuracy improves collective forecasting performance. Implementation may allow improved anticipation of and response to emerging risks and improved human collective efforts generally.

**Significance Statement:** Human-machine hybrid approaches have been identified as a new frontier for event prediction and decision making in the artificial intelligence and collective human intelligence fields. For the first time, we present the successful development and validation of a human-machine hybrid prediction market approach and demonstrate its superior accuracy when compared to prediction markets based on human forecasting alone. The advantages of this new hybrid system are demonstrated in the context of COVID-19-related event prediction.

## Introduction

Forecasting future events is an essential human capacity but reaching sufficiently high accuracy via human-only methods is a challenge. For example, the COVID-19 pandemic highlighted that traditional epidemic forecasting methods have performed poorly^1^. Such challenges pose problems for the future of decision making in an increasingly complex world. While decision making increasingly relies on artificial intelligence (AI), it is well known that the open-ended nature of many human problems is difficult for AI applications to capture^2^. These issues with task complexity in AI have led the field to develop the concept of human-machine hybrid approaches, particularly when human-only methods also falter^3-5^. This paper presents important first steps in progressing the collective intelligence application of prediction markets towards a hybrid model for better decision making across contexts.

Collective intelligence is the phenomenon whereby groups can outperform individuals when performing cognitive tasks^6^. This idea was rarely utilised in the COVID-19 crisis, partly because there is a paucity of evidence on how collective human thought should be elicited, collated, and analysed to reduce error^7^. Prediction markets are the typical application of collective human forecast systems^8,9^. In these platforms, individuals contribute inputs to group decision-making by trading on a continually updating market signal on the likelihood of future events^8^. Some studies indicate that the predictive performance of such markets may be higher than surveys, expert panels, and polls^10-12^. These systems allow cognitively diverse yet independent input from many people, a key feature of ‘wisdom of the crowd’ approaches^12^. The iterative aggregated market signal provides efficient member feedback^8^. Social network plasticity and feedback over time are key adaptive mechanisms for refining judgements and improving accuracy^13^. A market mechanism outperformed nine other ‘wisdom of the crowd’ aggregation mechanisms in recent controlled experiments^14^. Thus, prediction markets provide a promising base to develop better collective intelligence for forecasting future events.

The Next Generation Social Science program (NGS2)^15^ employed this system successfully, pooling 103 markets from four large-scale scientific replication projects across psychology^10^, economics^16^, and social science^17^. These prediction markets were correctly able to predict a higher proportion of replication outcomes than surveys^10,15^. Forecasters (traders) could choose to spend more points on trades that they were more confident would be accurate, providing a greater eventual point return. Thus, these active markets allowed a real-time weighting of the trade at execution by the active trader. The actual executed trades were then directly utilised for further investigations.

Further developments are now possible. Prediction market frameworks that support human traders and machine agents to make predictions could be developed. Such systems would allow human intelligence to be leveraged at scale over large amounts of data with the use of machine-based pre-processing or post-processing^4^. This would provide an anthropogenic artificial intelligence, where machine learning is applied to human cognitive outputs^5^. Some features previously associated with higher accuracy in prediction markets, such as small, frequent updates^18^, only emerge at the time of trade. In financial markets, Bayesian updating trading behaviour patterns distinguish those with higher accuracy^19^, an extreme example being distinguishing inside traders from noise traders. Other features such as past forecasting performance are generated outside the active market but are also likely important^20^. Machine learning may enhance our ability to harness these features. In a recent example in earthquake monitoring, human monitoring of the seismic signal time series was enhanced by machine learning processing of initial signals to remove anthropogenic noise, increasing the signal-to noise ratio^21^.

It is possible that further improvements to predictive performance could be accessed by rating trades for likely accuracy and then differentially weighting trades by these ratings in real-time. The validity of survey and other human report data can be improved by post-processing human data inputs by weighting responses^22^. This post-process weighting has not yet been applied to prediction markets, hindered by a complex, dynamic, trading microstructure.

Our approach improves the accuracy of collective forecasts from 1,878 markets from the Almanis platform^23^. External validation is undertaken using the scientific reproducibility program on the Next Generation Social Science (NGS2) platform^15^. In Phase 1, we used prediction market data to build a model for forecaster accuracy, validated it in two out-of-sample datasets, and demonstrated how human forecasts, when labelled with a higher machine-generated accuracy rating, had better predictive performance. In Phase 2, we applied a post-processing algorithm to weight human forecasts by their likely accuracy machine rating; this hybrid human-machine information system made more accurate predictions than prediction markets based on human forecasting alone. We then demonstrated the advantages of using this new hybrid approach specifically for COVID-19-related event prediction.

## Results

Across the two platforms (Fig. 1), all raw prediction markets (without optimisation using our processes) were significantly better than chance to forecast event occurrence (all; p<1×10^−6^ (Supplementary Table 1 and 2)). Consistent with past work, the prediction markets improved in accuracy as time passed, with higher AUCs as question settlement approached (Supplementary Table 3).

**Figure 1.**
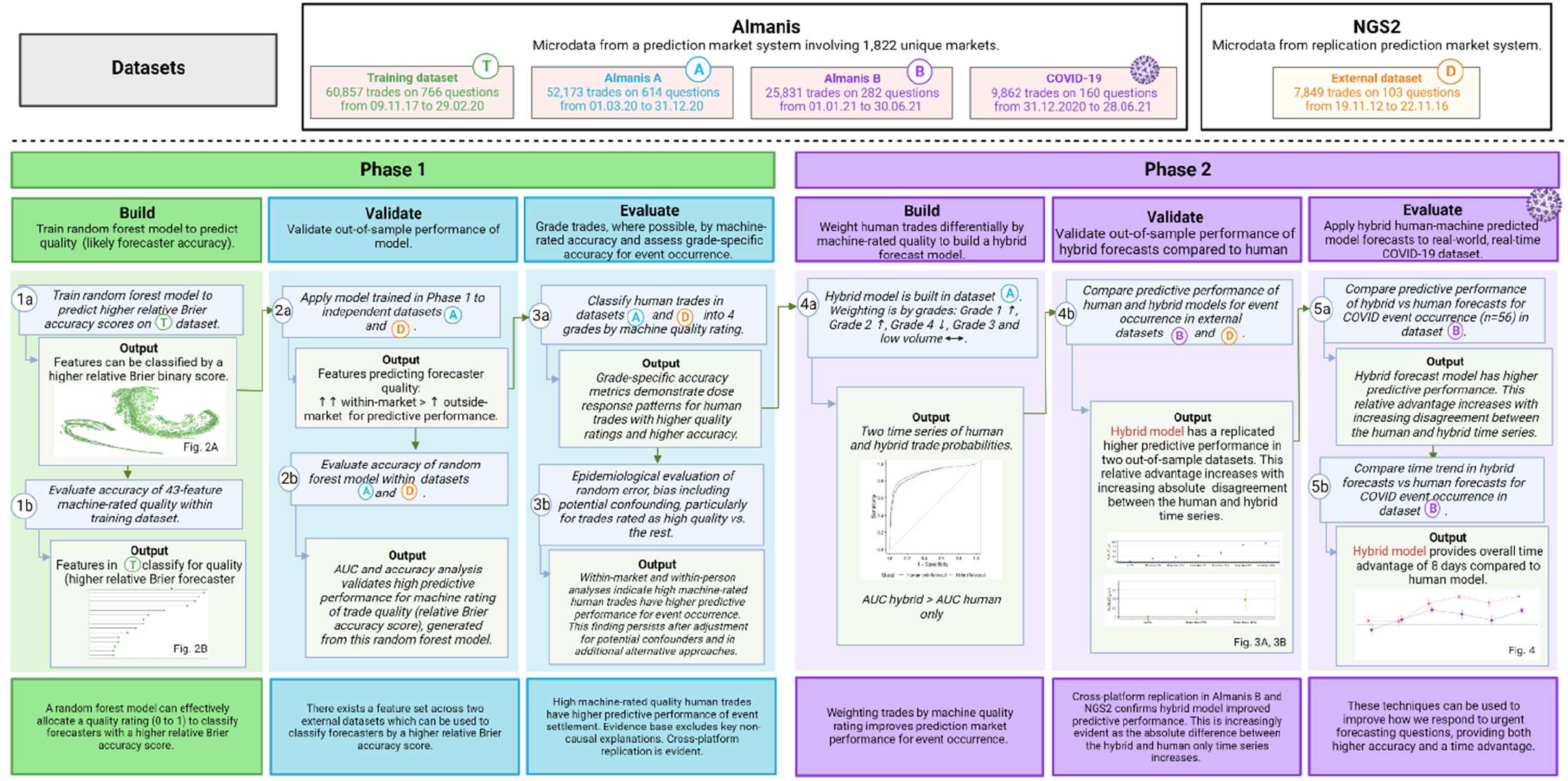
The hybrid information system of human collective forecasting with machine quality rating for event prediction.

### Phase 1. Building, validating, and using the forecaster accuracy model

Multiple features of the Almanis training set dataset classified forecaster accuracy, the top quintile of relative Brier scores, with good discrimination (Fig. 2A). A random forest model to predict this top quintile outcome was then developed. Ranking individual features by variable importance, the top ten were generated internally from forecaster behaviour in the market in question while the next five were related to their forecasting behaviour external to the given market, that is, in a previous or concurrent market (Fig. 2B). The full random forest model comprised of 43 variables provided each trade with a real-time continuous accuracy probability, from 0 to 1, of being in the top quintile relative Brier accuracy score. This is also termed ‘machine quality rating’. After a ten-fold cross validation, the full model had an AUC of 0.904 (95%CI 0.899, 0.909), p<1×10^−15^. We then validated the forecaster accuracy model, applied in real-time on each trade, on two out-of-sample datasets. This provided an AUC 0.839 (95%CI 0.831, 0.848) and 0.593 (95%CI 0.569, 0.618) for predicting trades in the top quintile of relative Brier accuracy in Almanis A and NGS2, respectively (Supplementary Fig. 1).

**Figure 2.**
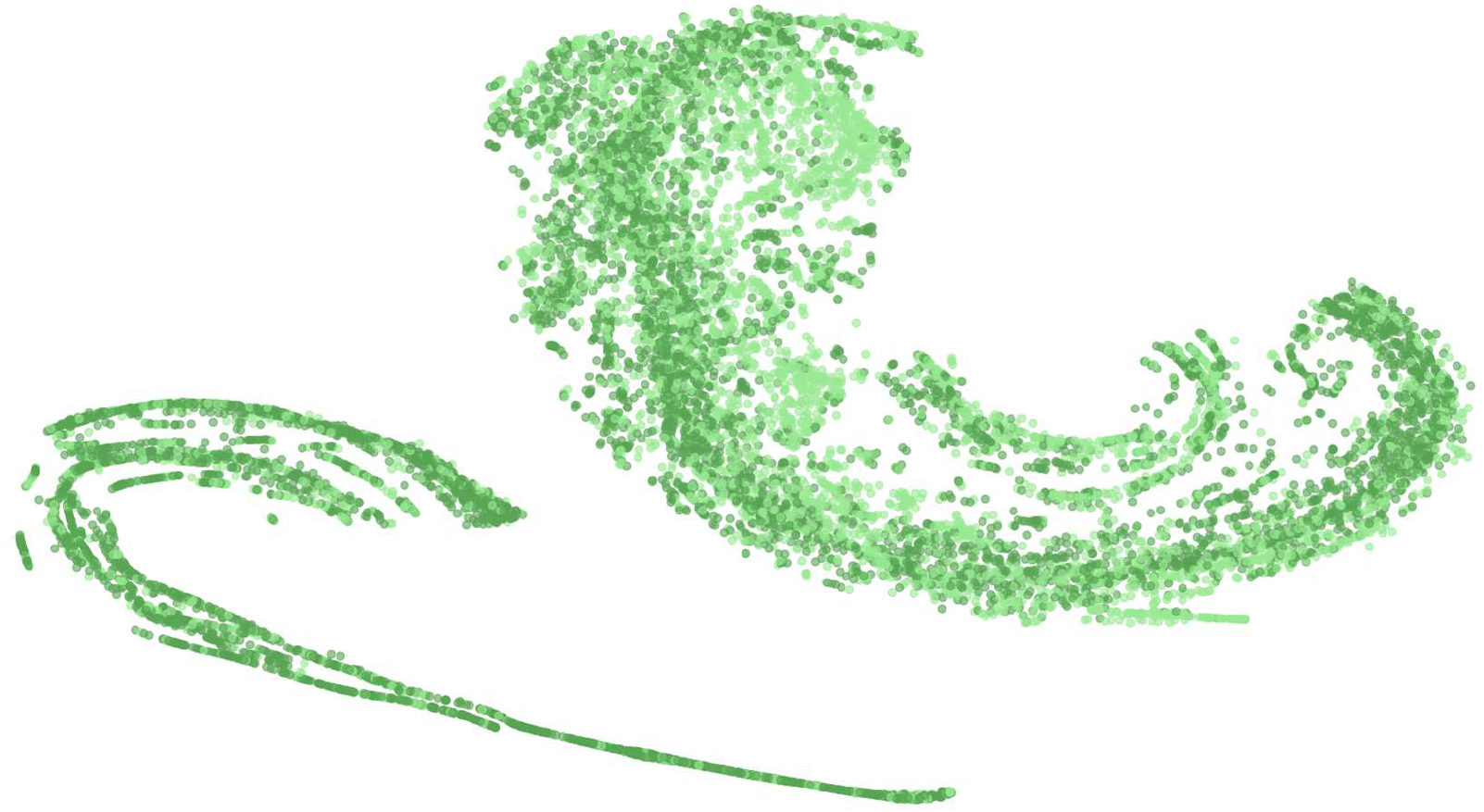
Development of a machine learning accuracy model that allows stratification of forecasts by likely forecaster accuracy. **Figure 2A: A t-SNE plot visualisation of classification by top quintile of forecaster relative Brier accuracy**. This figure indicates highly performing (top quintile for relative Brier accuracy probability) forecasters (dark green dot) vs. other forecasters (light green dot), based on the 43 features included in the random machine learning models. Dataset: Almanis training set with 766 prediction markets.

**Figure 2B:**
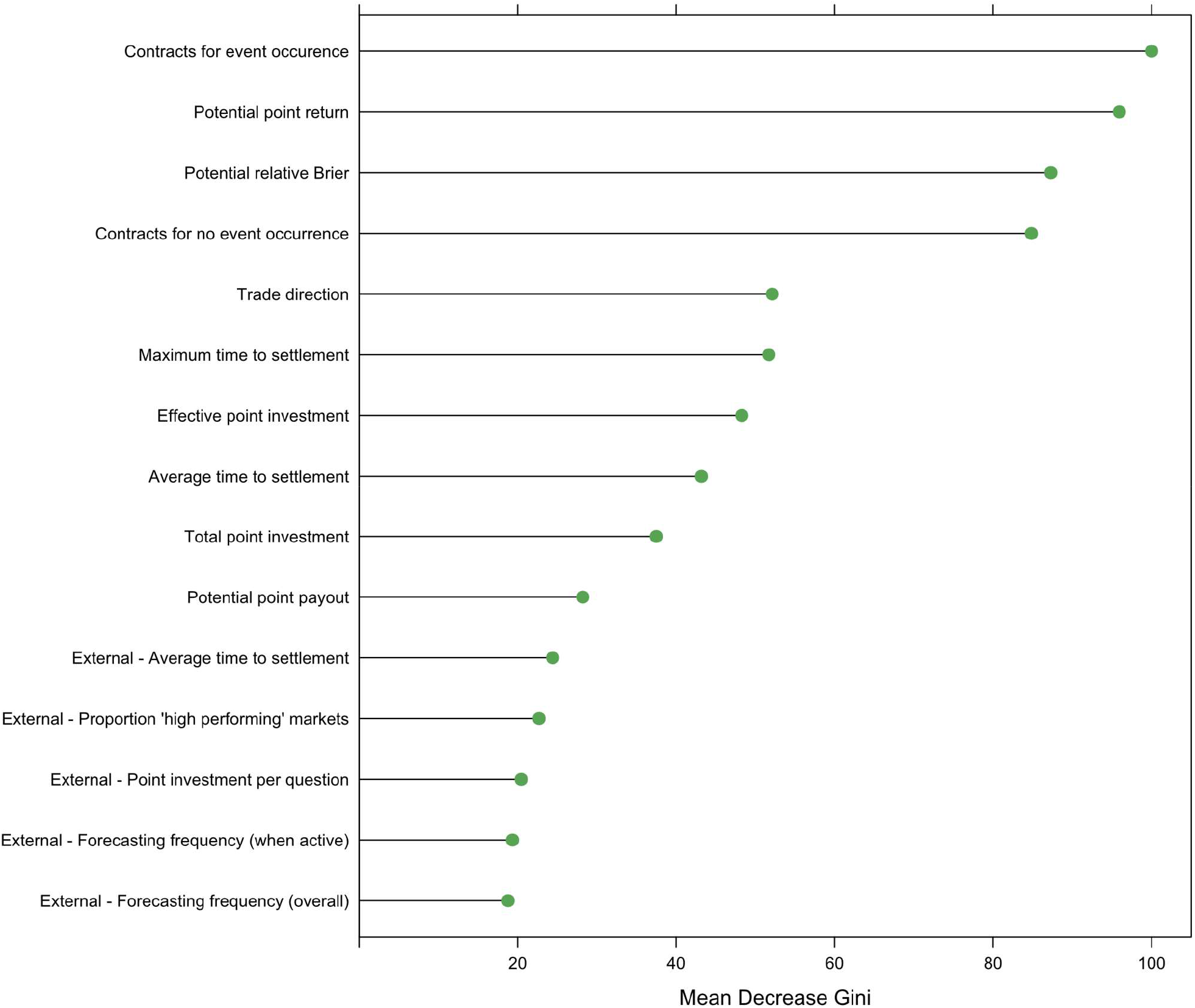
The top ranked features in the random forest model predicting whether a forecaster is in the top quintile for relative Brier accuracy probability: within-market features are more highly ranked than external features. All features except those marked as ‘External’ are based on a forecaster’s trades to date within a given question. Contracts for event occurrence, the number of ‘yes’ contracts a forecaster has bought (these contracts will return 1 point if the event occurs and nothing otherwise); Potential point return, potential points earnt (if forecasts are correct) minus points invested; Potential relative Brier, the maximum relative Brier possible supposing the best possible outcome of a forecaster’s forecasts; Contracts for no event occurrence, the number of ‘no’ contracts a forecaster has bought (these contracts will return 1 point if the event does not occur and nothing otherwise); Trade direction, the number of ‘yes’ responses by a forecaster as a proportion of their total number of forecasts (a yes response is a trade that will have a positive return if the event occurs); Maximum time to settlement, time from forecaster’s first trade to question settlement; Effective point investment, points invested in the question as well as how many points any ‘cashing out’ trades ‘would have cost’; Average time to settlement, the average time between a forecaster’s trades and question settlement; Total point investment, total amount of points invested in the question; Potential point pay out, potential points earnt if forecasts are correct; External – Average time to settlement, average time to settlement across forecaster’s trades on other questions; External – Proportion ‘high performing’ markets, proportion of markets (which forecaster traded on) where forecaster qualified as ‘high performing’ – i.e., had a relative Brier score placing them in the top 20% of forecasting performances in the training set; External – Point investment per question, the mean points invested by a forecaster on other questions they forecast on ; External-Forecasting frequency (when active) - forecasting frequency when a forecaster has an active position in a market; External - Forecasting frequency (overall) - forecasting frequency over the entire duration of a market. For external indices, the majority of information was sourced from past markets with a small contribution from concurrent markets. Dataset: Almanis training set with 766 prediction markets.

**Figure 2C:**
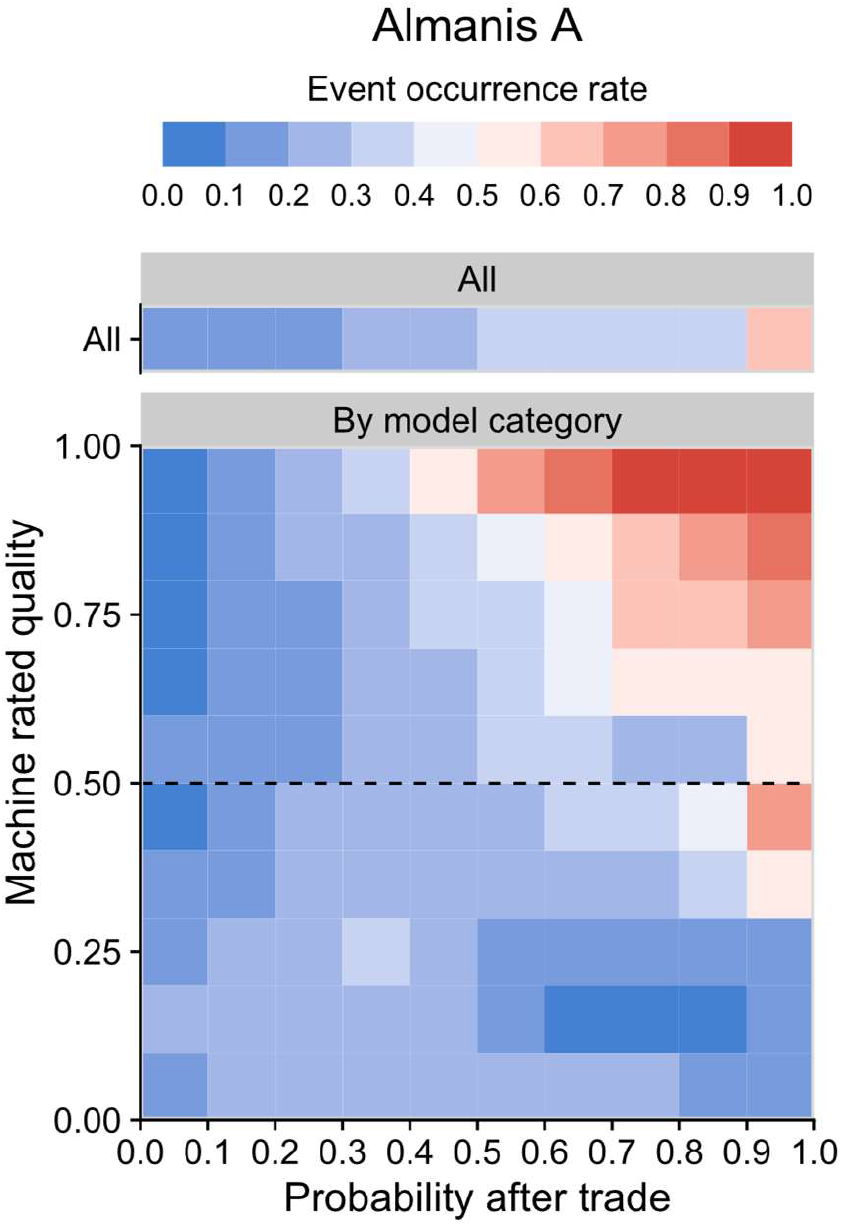
Human forecasts predict subsequent event occurrence rate: the agreement is potentiated as machine rated quality increases in Almanis A. Using multiple linear regression, outcome was the event occurrence rate defined as the proportion of markets where an event occurred of all markets. In the top plot, the ‘all’ samples are unstratified and adjusted for market-wide Brier error score, time to settlement and before trade probability. In the lower plot, the samples are stratified by grades of machine rating of trade quality and additionally adjusted for the interaction between before trade probability (0 to 1) and trade quality (0 to 1). Trade quality grades are displayed in 0.1 increments. There was only a weak correlation between human post-trade probability and machine quality rating (r =-0.14 (95% CI -0.15, -0.13)). In Almanis A, this is evident when one compares machine quality rating Grade 0 < 0.1, where regardless of the magnitude of the human forecast, the actual event rate is less than 0.5 (event occurred in < 50% of all markets) compared to machine quality rating Grade 0.9 ≤ 1.00, where the magnitude of the human forecasts matches the actual event occurrence rates very closely. The unstratified human forecast (All), as expected, provides an intermediate result between these two extremes. Overall, the trade probabilities predicted subsequent event occurrence rate and the agreement was magnified (p_*difference in effect*_<1×10^−15^) as trade likely accuracy quality increased. Dataset: Almanis A (n=614 markets).

Trades with a higher machine likely accuracy rating had post-trade probabilities which more closely resembled the actual event occurrence rate; for both Almanis A and NGS2, p*difference in effect*<1×10^−15^ (Fig. 2C). Consistent with this, the AUC gain across both Almanis A and NGS2 increased with higher grade trades (Supplementary Fig. 2; Supplementary Table 4). Forecasts graded as high quality (with an accuracy probability over 0.5) had a greater AUC gain vs other trades of lower quality (AUC gain per trade, Almanis A; 0.6 % (95%CI 0.4, 0.8%) vs. -0.3% (95%CI -0.6, -0.06%). The higher AUC associated with high quality trades occurred earlier from settlement than for other trades, providing a time gain advantage (Supplementary Table 5). The vast majority of forecasters, 91.7% (563/614), made at least one high quality trade; these forecasters still only had a high-quality trade in approximately half the markets in which they participated (Supplementary Fig. 3). For forecasters who traded in five or more markets, the median proportion (interquartile range) of a forecaster’s markets where a high-quality trade was detected was 0.54 (0.34, 0.72) and 0.50 (0.32,0.75) for Almanis A and NGS2, respectively. We examined the extent that these findings were independent of market-level or forecaster-level effects in Almanis A. Using hierarchical logistic models to cluster by market, forecaster, or both, the probability of event occurrence was significantly higher (p<1×10^−16^, <1×10^−4^ and <1×10^−4^) for high quality forecasts compared to other trades.

**Figure 3.**
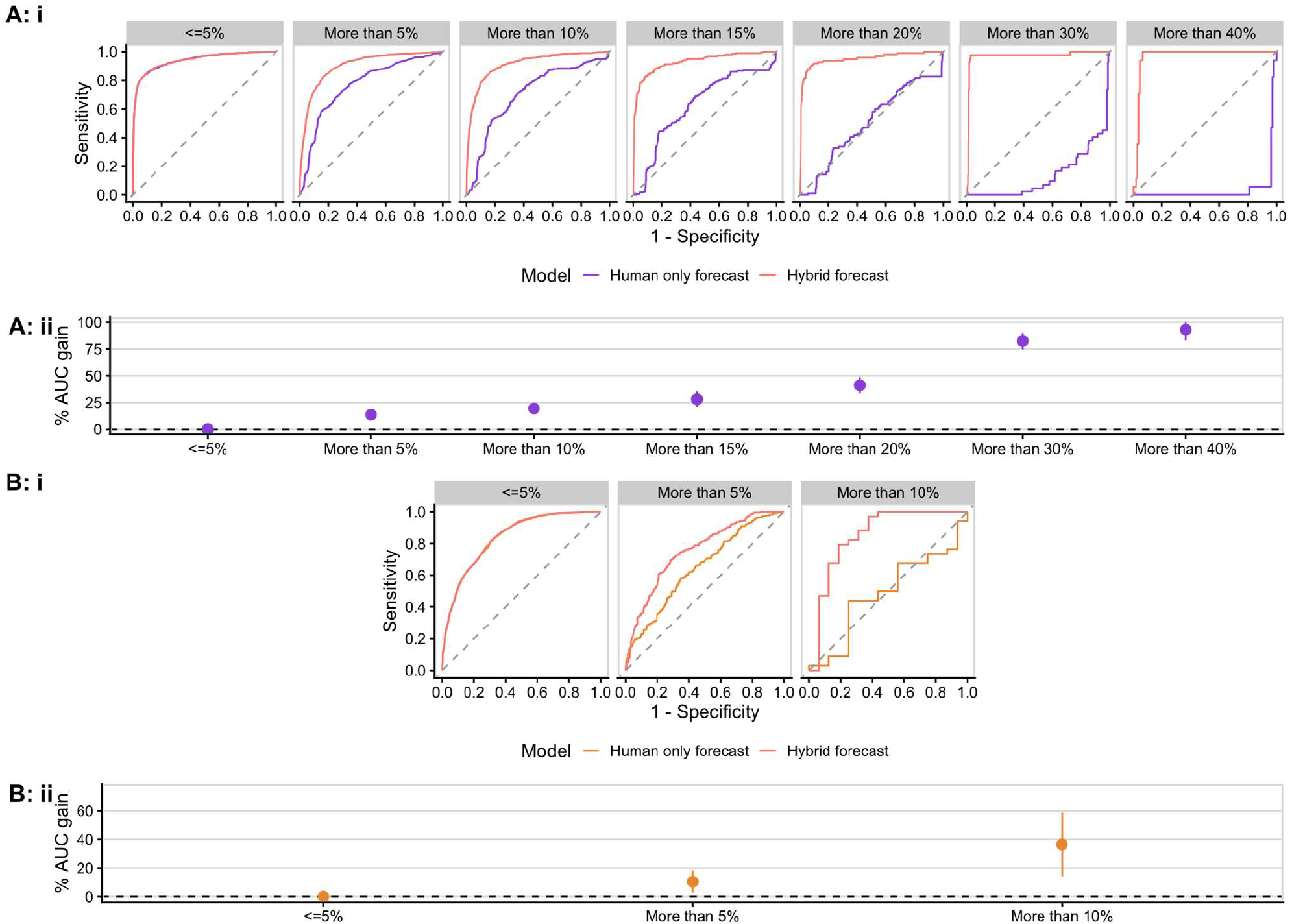
The predictive performance advantage of the hybrid vs. human only model increases with higher disagreement levels. i. Area Under the Curve, and ii. Temporal Area Under the Curve gain (95%CI) due to an active trade. for hybrid vs. human only forecast model: stratified by disagreement level at the time of trade. Fig. 3A: Almanis B. i. Human only vs hybrid AUC (p value of difference, clustered in market). All (100% of trades): *0.915 vs 0.931, p-value=6.104×10*^*-9*^. ≤ 5% (80.9% of trades): 0.940 vs 0.940, p-value=0.509. More than 5% (19.9% of trades): *0.768 vs 0.904, p-value=1.35×10*^*-14*^. More than 10% (12.5% of trades*): 0.707 vs 0.913, p-value=<1.00 × 10*^*-15*^. More than 15% (8.1% of trades): *0.6429 vs 0.930, p-value<1×10*^*-15*^. More than 20% (5.7% of trades): *0.502 vs 0.950, p-value=<1.00 × 10*^*-15*^. More than 30% (2.2%): *0.137 vs 0.969, p-value<1×10*^*-15*^. More than 40% (0.8% of trades): *0.039 vs 0.964, p-value<1×10*^*-15*^. **ii. AUC gain (hybrid – human only) (95%CI)**. Overall: 1.6 (1.06, 2.14). ≤ 5%: 0.03 (−0.07, 0.14). More than 5%: 13.21 (9.85, 16.58). More than 10%: 20.57 (16.42, 24.72). More than 15%: 30.09 (23.60, 36.60). More than 20%: 44.76 (37.61, 51.90). More than 30%: 83.26 (76.83, 89.69). More than 40%: 92.44 (81.04, 100). **Definitions:** disagreement is the % absolute difference in after trade probability between the human only and hybrid forecasts. I.e., the 5% category shows trades which differ between human only and hybrid forecasts by more than ±5% probability. Dataset: Almanis B (n=282markets). **Figure 3B: NGS2. i. Human only vs hybrid AUC (p value of difference, clustered in market)**. All (100% of trades): 0.827 vs 0.833, p-value=0.097. <=5% (82.4% of trades): 0.837 vs 0.838, p-value=0.972. More than 5% (17.6% of trades): *0.649 vs 0.755, p-value=0.008*. More than 10% (10.8% of trades): *0.485 vs 0.851, p-value=0.001*. **ii. AUC gain (hybrid – human only) (95%CI)**. Overall: 1×10^−2^ (−0.30, 0.31). ≤ 5%: 10.5 (2.75, 18.3). More than 5%: 36.58 (14.32, 58.84). More than 10%: 0.62 (−0.11, 1.34). **Definitions:** disagreement is the % absolute difference in after trade probability between the human only and hybrid forecasts. Dataset: NGS2(n=103markets).

### Phase 2. Improved prediction of event occurrence is evident for human trade probabilities after weighting by machine quality rating: a hybrid model

We built a hybrid event prediction model using Almanis A which differentially weighted trades by machine-derived accuracy probability in real-time. This hybrid model had a higher AUC than the human only model and this was evidenced not only in the test set (p=3.791×10^−10^) but at out-of-sample Almanis B and NGS2 (Fig. 3, Supplementary Table 6). The better relative performance of the hybrid model compared to the human only (general) model increased as the two forecast series disagreed more strongly. When the two models disagreed by 5% or less, their predictive performance was similar (Fig. 3). At more than 5% disagreement, the AUC gain for the hybrid vs. human model in Almanis B and NGS2 was considerable: 13.2% (95%CI 9.9%, 16.6%) and 13.8% (95%CI 4.6%, 23.0%) (Fig. 3). These findings were robust following further exclusion of any reliance on data sourced from external markets or related to time to settlement (data not shown) and remained evident in markets with 30 or less traders with an AUC gain of 12.4% (95%CI 7.9 %, 17.0%) and 10.5% (95%CI 2.8%, 18.3%) respectively. We examined the situation where the hybrid and general models disagreed on the likeliest event outcome (i.e., when they were on either side of the 0.5 threshold). This occurred on 528 trades across 84 markets. For these, the human market was correct for 22.6% (19/84) and the human-machine model was correct for 77.4% (65/84) of these markets; p=2.4×10^−7^. Thus, the net reclassification benefit from using the hybrid rather than human only model was marked in Almanis B.

We then applied this method to the COVID-19 event markets (Fig. 1). Overall, the hybrid model had higher accuracy than the human model (p=0.026), with an AUC gain of 0.38% (95%CI 0.05%, 0.71%). The differences in mean absolute accuracy score, an indicator of the closeness of the distance between post-trade probability and true event occurrence, become increasingly evident with increasing disagreement from more than 5%, 10% and 15% disagreement, pall<1×10^−4^. Fig. 4 shows the increase in mean absolute accuracy score in the COVID-19 markets as time to settlement shortens stratified by 10% disagreement or not. Thus, given that markets become more accurate towards settlement, the hybrid model also provides a relative time advantage for forecasting.

**Figure 4.**
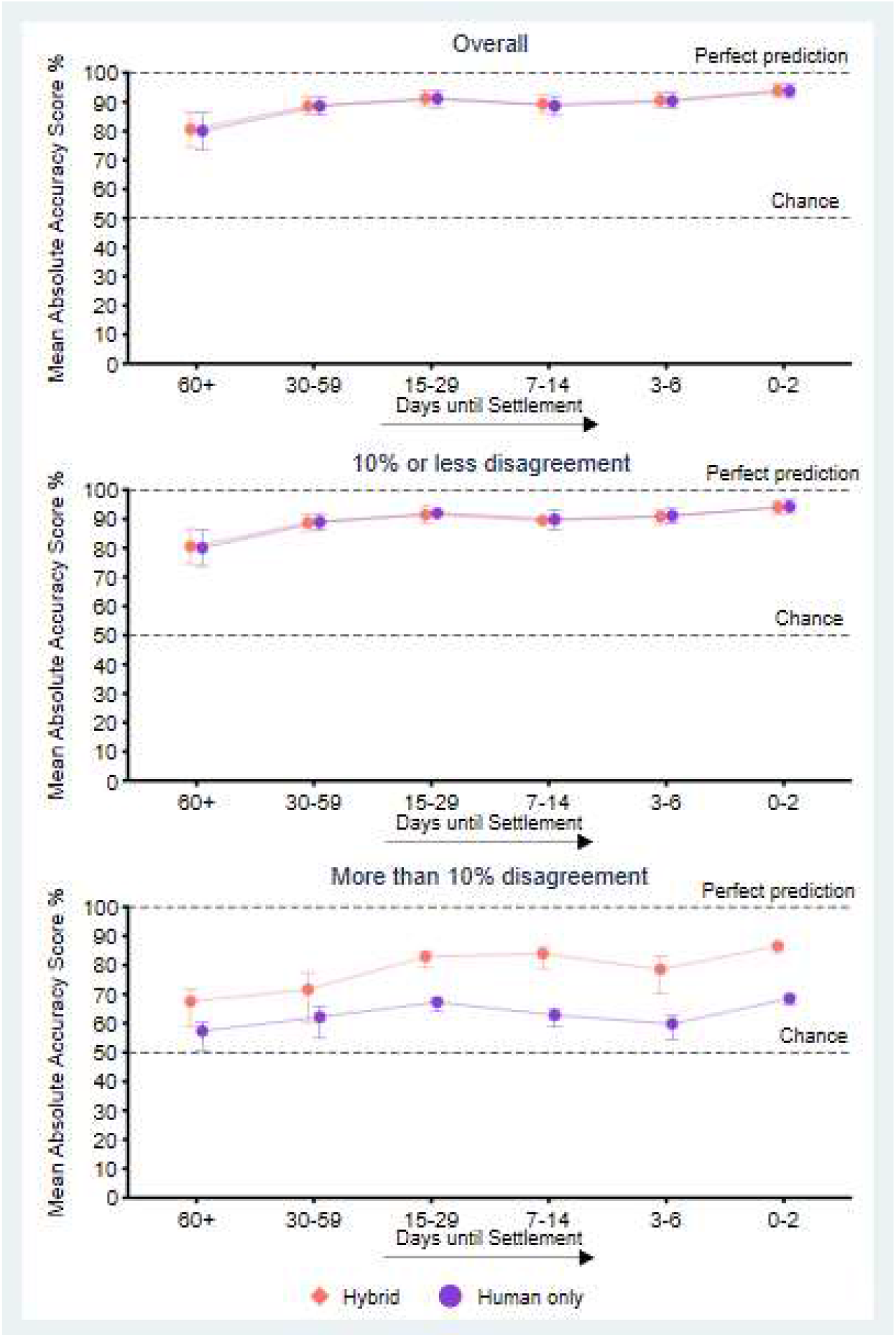
The accuracy improvement of hybrid vs human only market for COVID-19 event prediction provides a time lead for accuracy towards settlement. The higher accuracy of market signals towards settlement (Supplementary Table 3), combined with the greater accuracy of the hybrid model seen here, provides a time lead in accuracy when compared to the human only market signal. Here, the level of agreement to the binary COVID-19 event outcome (1=event occurred, 0=event did not occur) is provided by the mean absolute accuracy score, 1-MAPE, where 100% would indicate perfect prediction, 50% the prediction expected by chance (a coin toss) and 0% complete disagreement. Here, when the human only and hybrid forecasts have 10% or less disagreement, the performance of the two series are equivalent. When the disagreement between these is more than 10%, then the hybrid market is more accurate at a given time point. Dataset: COVID-19 markets (n=160).

On a practical level, government responses to COVID-19, such as community lockdowns, are often based on the assumption that an event, such as hospitalisation numbers exceeding a threshold, will or will not occur. Therefore, we examined the situation where the hybrid or general model was discordant (opposing) for event outcome. This occurred in 33 markets involving 171 trades. For these, the human market was correct for 27.3% (7/33) and the human-machine model was correct 72.7% (26/33) p=0.007. This finding replicates the net classification improvement obtained by using the hybrid model similarly in the Almanis B sample p=2.4×10^−7^. In both datasets, the net classification improvement of using the hybrid rather that human forecast when forecasts were discordant was higher in markets with higher divergence between the hybrid and general market (Supplementary Table 8).

## Discussion

Prediction markets are a collective intelligence system based on human forecasts. Here, we created a hybrid information system, incorporating human-sourced trades and real-time machine trade quality ratings, to generate more accurate hybrid human-machine forecasts. Where the human-machine hybrid and the human only forecast model disagreed by 5% or more, the hybrid forecast accuracy advantage was substantial, corresponding to highly significant 13% and 14% gains in AUC in the out-of-sample data sets of Almanis B and NGS2, respectively. We demonstrate applicability to COVID-19 events: -when the human-machine and human model disagreed on whether a COVID-19-related event would occur or not, the hybrid model provided the correct prediction 73% of the time. The majority of the analyses were conducted at the trade level, providing highly significant findings. Further, the findings from Almanis were validated externally in NGS2 scientific replication studies and/or in subsequent Almanis samples.

The value of the hybrid market signal became most evident when it diverged strongly from the human only signal in the post-trade probability assigned to event occurrence. Although the two signals were similar after the majority of trades on the Almanis platform, more than one tenth of trades led to the two signals diverging by 10% or more difference in forecast event probability, where the hybrid forecast accuracy advantage was highly significant. On a practical level, one would be particularly likely to rely on the hybrid model when making assumptions about whether an event will occur if it disagrees strongly with the human only signal. Where the two systems disagreed on whether the event would occur or not, the hybrid signal was correct 77% and 73% of the time in Almanis B and COVID-19, respectively. This is significantly greater than chance, which would have seen each system correct roughly at 50% of the time.

Every day, people make countless decisions based on the likelihood of future events. Here, the findings further improve the value of prediction markets over other forecast methods. The model to predict forecaster accuracy was validated in two out of training sample datasets with within-market features and external features important predictors. The contribution of within-market features was less evident in NGS2, where trading activity mainly occurred in the first week and the relevant information was available at market open.^15^ Some behavioural features we observed have been reported previously, including higher trade activity, small and frequent trade updating, and changing trade direction.^18,19^ Some of the external features of past accuracy and past return on point investment are consistent with the notion of Superforecasters® previously identified, in part, on cognitive abilities.^20^ However, high quality trades were not evident in forecasters as a fixed trait - most forecasters made at least one high quality trade, but only about half of their trades were high quality. Trades graded as high likely accuracy quality were more accurate than other trades even by the same forecaster in the same market. There was an increasing superiority of performance by the hybrid forecast model as disagreement between the human-machine hybrid and human only model increased.

Here, the creation of a likely accuracy quality indicator at the time of the forecast input, combined with the prioritisation of forecasts determined to be more likely to be accurate, is shown to improve the signal-to-noise ratio by reducing prediction error. These findings indicate a way to advance internet-based human collective intelligence. Currently, most human crowdsourcing inputs, such as those on social media, have no concurrent signal on likely importance. In the natural world, prioritisation of individual inputs to the collective is evident in herds^24^ and swarms.^25^ The individual agents can provide, in addition to signal information, a concurrent signal rating, often transmitted through a real-time behaviour pattern,^25^ to the collective. The collective then responds differently to the input depending on the accompanying rating for threat^24^ or potential reward.^25^

The two prediction market platforms shared useful design features including both operating under the logarithmic market scoring rule with a point system to incentivise accurate forecasts. The forecast probabilities and the quality ratings metrics were only weakly correlated. This is consistent with recent recommendations to keep humans and machine measures independent, if possible, to leverage hybrid performance.^26^ Selection forces did appear to be occurring to some extent in the NGS2 dataset with high quality trades more likely to be made in less certain markets. We used various approaches to account for this, such as reporting the AUC gain after vs. immediately before an instantaneous trade. Overall, we employed an approach to reduce or evaluate non-causal explanations such as confounding or bias.^22,27^ We either standardised for, or assessed, factors varying:- (i) at the market-level (e.g. market size or topic), (ii) intra-market variability (e.g. accuracy improves towards settlement date^28^) and (iii) inter-personal variability (e.g. forecaster ability^20^) by using a variety of approaches. These included hierarchical models with market, time, or forecaster set as a random effect. We conclude confounding or other bias are unlikely to explain these results. Our approach triangulates causal features beyond replication alone.^22,27^ There is some analogy with the ‘surprisingly popular’ concept, an indicator of wisdom in the crowd,^29^ because the high machine quality rating trade profiles became popular, particularly within a week of market close, indicating that higher quality trades anticipated the market.

This work has direct application in a range of settings where it is important to either improve forecast accuracy or improve the speed with which one arrives at a given forecast accuracy, or both. We demonstrated one potential translational application: an improved predictive accuracy for COVID-19 events. The use of prediction market information systems has already been steadily increasing^9^ as (i) evidence accumulates on their superior forecasting performance and (ii) disruptions to environmental and human systems with new associated risks mean that forecasting models built on historical inputs only are less reliable than newer systems that harness dynamic human collective intelligence. We anticipate the further development of hybrid human-machine information forecasting systems. In addition, these findings have broader application to other current human crowd sourcing platforms, where rating and treating inputs differentially by their value may improve the signal-to-noise ratio.

## Materials and Methods

### The Almanis prediction market platform

Almanis is a prediction market across geopolitical, economic, health and other domains.^23^ Each player, enrolled through global social media, was given an account with an operator-staked set of 1,000 finite points with point gain (with financial reward and prizes) or loss based on their contribution to moving the market towards or away from correct event occurrence prediction (Supplementary Methods). An automated market maker using the logarithmic market scoring rule (LMSR) was used to facilitate trades.^30^ The emerging price for the contracts traded in the market can be interpreted as the aggregated expected probability of event occurrence. Each active market generated a large variable set across trade-level (e.g., the frequency and timing of trades), forecaster-level (e.g., past performance) and the market-level (e.g., question topic) (Supplementary Methods). No personal identifying information was collected. Ethical approval for this longitudinal project was obtaining from the Royal Children’s Hospital Human Research Ethics Committee, Melbourne, Australia (2018, 38248). We report on several mutually exclusive prediction market samples (n=1,822 in total) from the Almanis platform (Fig. 1).

### Next Generation Social Science scientific replication prediction markets

This pooled dataset consisted of the Reproducibility Project: Psychology;^10^ the Experimental Economics Replication Project;^16^ the Many Labs 2 project;^31^ and the Social Science Replication Project.^17^ Each of the four had a focus on forecasting the likelihood that scientific study findings would be replicated. People working in academia were recruited via social media and mailing lists. The trading system was also an operator-staked LMSR platform with an inital token allocation then gains converted to monetary rewards.^15^ A key design difference to Almanis was that forecasters in NGS2 were expected to forecast based on information provided only at market open (the key finding publication and the planned replication study protocol (Supplementary Methods)). This longitudinal dataset of 103 markets (Fig. 1) was used for external validation of the Almanis findings.

### Statistical methods

The Almanis sample size, even for COVID-19-related markets alone, had more than 95% power to detect a correlation of 0.58 between market likelihood (price) and binary settlement (0/1) at alpha=0.01. Adequate statistical power has been reported for the pooled NGS2 validation set.^15^ We employed an epidemiological longitudinal analysis approach.^22,27^

### Phase 1. Forecaster accuracy model

Random forest machine learning was used across multiple features in the 768 markets in the Almanis training set to predict if a forecaster had a top quintile relative Brier accuracy score^32^ (Fig. 1). A random forest model with 500 decision trees was trained using 10-fold cross-validation repeated 10 times (Supplementary Methods). The random forest model allowed the correlated features to be examined without allocating importance to one feature only within the correlated set.^33^ The algorithm randomly sampled a time point in the trade time series of each forecaster. Area under the curve (AUC) and other evaluation metrics are provided (Supplementary Fig. 1). The mean decrease in the Gini index^33^ was used to rank the final selected features for importance (Fig. 2B).

Independent validation of the random forest’s ability to identify accurate forecasters was conducted in Almanis A (614 markets, not used for training) and the external NGS2 platform (Fig. 1). The model assigned a machine likely accuracy rating to the trade series performance by a forecaster. This rating corresponded to the probability that the performance would earn a top quintile relative Brier accuracy score. Receiver operating characteristic (ROC) analysis^34^ was performed (Supplementary Fig. 1). We then assessed how event prediction performance varied by forecaster accuracy rating using several methods (Supplementary Methods). We graded forecasts in Almanis A and NGS2 by the machine accuracy rating of each selected trade into four grades (Supplementary Methods), terming those in the top two grades as high-quality trades. We report the AUC gain after vs. immediately before an instantaneous trade^35^. AUC between different models on the same sample or across independent samples were compared by using the approach of Obuchowski et al.^36^ to cluster within market to reduce the influence of any market-level effects and account for the correlated nature of forecasts within the same market.

### Phase 2. Event prediction using a hybrid human-machine model

We built a hybrid machine-human predictive model based on human forecasts then weighted by machine accuracy probability. In brief, the model was built using cross-validated grid search to optimise parameters on the Almanis A dataset (Supplementary Methods). Validation was then undertaken on the independent Almanis B and NGS2 datasets (Fig. 1). Clustered AUC analysis^36^ and a hierarchical regression model were used to allow intra-market comparisons. We then stratified by the level of disagreement between the hybrid model and the general market at the time of trade. We examined a further independent sample of COVID-19 related markets using various methods (Supplementary Methods). These markets forecast on likely infection rates, emergent COVID-19 strains, vaccination rates, and likely pandemic policy responses as listed in the Supplementary Table 7. In total, 434 forecasters made 9,862 trades across 160 COVID-19 event markets. All computations were performed in R code,^37^ STATA 16, or Python.

## Supporting information

Supplementary Methods and Results

## Data Availability

Data availability. The dataset from Almanis prediction markets was used under license for the current study and is not publicly available. Applications for access to the Almanis database can be made from Dysrupt Labs, Slowvoice Pty. Ltd. NGS2data can be accessed through an R package found at: https://github.com/MichaelbGordon/pooledmaRket.
Code availability. Data collection: Provision and use of the code for the Almanis platform is subject to a Commercial License Agreement with Dysrupt Labs, Slowvoice Pty. Ltd. Data analysis: Provision and use of the code is generally subject to a Commercial License Agreement with Dysrupt Labs, Slowvoice Pty. Ltd. For academic and government purposes, please contact the corresponding author. All experiments and implementation details are described in sufficient detail in the appendix to support replication with non-proprietary libraries.

## Data and materials availability

### Data availability

The dataset from Almanis prediction markets was used under license for the current study and is not publicly available. Applications for access to the Almanis database can be made from Dysrupt <u>Labs, Slowvoice Pty. Ltd. (</u>). NGS2data can be accessed through an R package found at: https://github.com/MichaelbGordon/pooledmaRket.

### Code availability

*Data collection*: Provision and use of the code for the Almanis platform is subject to a Commercial License Agreement with Dysrupt Labs, Slowvoice Pty. Ltd. *Data analysis:* Provision and use of the code is generally subject to a Commercial License Agreement with Dysrupt <u>Labs, Slowvoice Pty. Ltd. (</u>). For academic and government purposes, please contact the corresponding author. All experiments and implementation details are described in sufficient detail in the appendix to support replication with non-proprietary libraries.

## Acknowledgments

This work was supported by an AusIndustry R and D tax incentive program from the Department of Industry, Science, Energy and Resources to SlowVoice Pty Ltd. (IR2101990) and an Investigator grant (N1197234) and Fellowship to A-L Ponsonby by the National Health and Medical Research Council of Australia (GNT1110200). We thank the Defense Advanced Research Projects Agency for providing open access prediction market datasets. We thank Cameron Patrick, Statistical Consulting Unit, University of Melbourne for statistical and data visualization advice. We thank Nitin Yadav, Brain, Mind & Markets Laboratory, University of Melbourne for discussions on concepts in this manuscript.

## Author Contributions

Conceptualisation: AG, KM, EM, FB, MC, CN, JI, ALP. Methodology: AG, KM, EM, FB, MC, CN, JI, ALP. Investigation: All. Visualisation: AG, EM. Funding acquisition: KM, FB. Writing – original draft: AG, KM, EM, MC, CN, JI, ALP. Writing – review & editing: All

