## Supplementary Methods and Results for "Machine learning augmentation reduces prediction error in collective forecasting: development and validation across prediction markets"

**This PDF file includes:**

Supplementary Methods
Tables S1 to S8
Figures S1 to S3
SI References

### Supplementary Methods

#### The Almanis prediction market platform

Almanis is a global prediction market run by Dysrupt Labs Pty Ltd.[1] It has a crowd of over a thousand active forecasters who make predictions via the market on the likelihood of various future events. The topics range across geopolitical, macroeconomic, scientific, health, technological, and financial domains.

#### Participants

Global recruitment commenced 1st December 2015 through English language Twitter and other social media globally. The forecaster distribution by region ranked as Africa, Asia, Europe, North America, Oceania, South America, and unknown.

#### Exclusions

Exclusion was based on failure to comply with Terms and Conditions, as agreed to at market entry. The most common non-compliance issues were forecasters operating multiple accounts or evidence of collusion with other forecasters.

#### The trading system

At market commencement, a question was displayed with information on the length of the market and the methods and sources that could be used to verify event occurrence or not at baseline. Where relevant, hyperlinks to relevant support material were also provided and it was expected that new relevant information would arise during the active market. Potential participants were made aware of operator-staked financial, reputational, and prize rewards based on forecasting accuracy and activity. Specifically, traders were allocated 1000 points in an account to purchase positions in markets of their choosing, and either lost points, eventually leaving the platform, or gained points which could be used in another market. They were rewarded with cash prizes based on their contribution to market movement towards correct settlement (event occurred or not) compared to other traders. The emerging price for the contracts traded in the market can be interpreted as the aggregated probability of event occurrence. Costly signalling was employed, a technique previously shown to improve accuracy.[2] The incentives payout over the Almanis-training study period between 2017 and 2020 amounted to £72,000, paid in cash through PayPal. T here were monthly pools of 4,500 Great British Pounds (GBP) that were generally distributed as 600, 300, 200, 100 GBP for the 1st, 2nd, 3rd, and 4th most accurate forecaster respectively, then lower increments for the remainder of the top 20 forecasters with a minimum reward of 10 GBP. However, this varied across the markets. Reputational and participation prizes were also awarded to maintain engagement.

#### Market maker

The logarithmic market scoring rule (LMSR) provided a buying and selling price at all times as a continual counter-trader.[3] The LMSR is further outlined in Equation 1 below.

#### Event occurrence

Markets are settled according to an event occurrence at market end coded as 1 or 0 if the event does or does not occur. This is reported by a verifiable source or range of possible sources, as stated at the opening of each market. Examples of the questions on binary event occurrence for the COVID-19 pandemic are listed in Supplementary Table 5.

#### Generated study measures

Using the crowd management system, Intercom,[4] information on connection times, web session number, browser type, and user location was collected. Variables were collected at the individual trade level (time, magnitude, direction, points allocated), the forecaster level (trade balance, past performance) and market level (e.g., question topic, question type (multiple-choice or binary), market duration). In addition, we created new features to represent time-based behaviour patterns informed in part by past human[5] and natural setting studies.[6,7] This included recording short trade series of up to four consecutive trades. This report is predominantly based on binary questions only except for the use of multiple-choice questions also for COVID-19. For the later, one response of the four was randomly selected per question, providing a binary outcome for whether this event occurred or not. The emerging price for the contracts traded in the market can be interpreted as the aggregated probability of event occurrence. A comment history was also recorded (users are able to comment free-text responses as part of markets). A more detailed variable list is provided in the table below, where the variables are classified as those varying within or external to a given market. This trading microstructure provides the features to identify high performing traders and generate collective forecasts.

**Ethics**
No personal identifying information was collected. Ethical approval for this project was obtaining from the Royal Children's Hospital Human Research Ethics Committee, Melbourne, Australia (2018 38248).

**Variables original from prediction market**
The Almanis dataset included the following variables for each trade:

| **Variable Name** | **Description** |
|---|---|
| QuestionName | Name of question |
| ProbBeforeTrade | Probability that market assigned to event occurring (i.e., a 'yes' outcome) before the trade |
| ProbAfterTrade | Probability that market assigned to event occurring after the trade |
| TradeTime | Date and time of trade |
| SettledValue | Settlement value for the question (0 or 1) |
| UserID | Identifier of user who made the trade |
| TimeToSettlement | Time until the question settled |
| AmountPurchasedYes | Number of 'yes' contracts purchased |
| AmountPurchasedNo | Number of 'no' contracts purchased |
| PointCost | Point cost of the trade (negative if trade involved closing out a position) |

**Variables Added to the Data**
A number of new variables were added to describe trader behaviour and performance. These were:

| Variable Name | Description |
|---|---|
| RelativeBrier | The relative Brier score of the trade (the calculation of this score is further explained in Methods) |
| AbsoluteROI | The point return on investment for each trade (this is calculated according to points earned minus points invested) |
| CumTradesOnQ | The cumulative number of trades on that question that the forecaster has made. (This is taken after the trade in question - so for their first trade, CumTradesOnQ = 1) |
| Answer | The outcome predicted by the forecaster (yes or no) |
| CumPosition | The forecaster's cumulative position in the question in terms of yes and no contracts. Cumulative position is calculated by subtracting a forecaster's no contracts from their yes contracts (so negative cumulative positions correspond to positions where the forecaster stands to benefit from a no outcome) |
| ForecasterUpdate | A variable indicating whether the trade reflected an update to the forecaster's belief (i.e., is not their first trade) |
| ForecasterUpdateSize | The amount that a forecaster updated their previous belief (NA for a user's first forecast) |
| ConsecutiveSameDirection | An indicator variable for consecutive trades a user makes in the same direction |
| nSameDirection for 2<n<5 | An indicator variable for n consecutive trades a user makes in the same direction |
| OppositeDirection | An indicator variable for a trade a user makes in the opposite direction to their previous trade |

These variables were either introduced to assess accuracy (RelativeBrier, AbsoluteROI) or to describe user behaviours which may be indicative of accuracy (all other variables).

**Creation of a Summary Table - Summarising by Question and UserID**

To assess the performance of each user on each question and identify user behaviours which were correlated with forecasting accuracy, a summary table was created. This included a number of variables describing user accuracy and behaviour, for each of the users who made at least one forecast. Each row in this summary table represented a user's performance on one question.

Examples of the component features of the full model (combined M1 + M2) random forest model for predicting informative trades in the top quintile of relative Brier accuracy values in Almanis-training and mean decrease in the Gini index are provided below.

| Variable | Mean Decrease Gini | Type[a] | Category | Description |
| --- | --- | --- | --- | --- |
| **TrueContracts** | 634.8171576 | Int | Size of investment/payoff | Number of True contracts bought in the course of the question |
| **PotentialMaxProfit** | 609.0722145 | Int | Size of investment/payoff | PotentialMaxPayoff' minus 'TotalCost' |
| **PotentialMaxRelativeBrier** | 554.4528387 | Int | Size of investment/payoff | The maximum relative Brier the forecaster could get supposing the best possible outcome of their forecasts |
| **FalseContracts** | 538.8346204 | Int | Size of investment/payoff | Number of False contracts bought in the course of the question |
| **Answer** | 331.9014774 | Int | Proportion of answer in same direction/repeated trades in same direction | Variable reflecting the proportion of 'true' responses by a forecaster |
| **MaxTimeToSettlement** | 329.2463199 | Int | Time to settlement | Time from forecaster's first trade to question settlement |
| **RealCost** | 307.6467196 | Int | Size of investment/payoff | Points invseted in the question as well as how many points any 'cashing out' trades 'would have cost' |
| **AverageTimeToSettlement** | 275.4111961 | Int | Time to settlement | Average time to settlement across all the forecaster's trades |
| **TotalCost** | 239.256648 | Int | Size of investment/payoff | Total amount of points invested in the question |
| **PotentialPayoff** | 180.672205 | Int | Size of investment/payoff | Maximum of 'TrueContracts' and 'FalseContracts' |
| **fprevavgTimeLeft** | 156.6605496 | Ext | Previous forecasting behaviours | Mean 'MaxTimeToSettlement' over previous questions |
| **fprevpropMarketsSignificantBrier** | 145.7742675 | Ext | Previous forecasting outcomes (how successful were previous forecasts) | Previous proportion of markets where user qualified as 'high performing' - i.e., top 20% of forecasting performances in the training set |
| **fprevcostPerQ** | 131.4451307 | Ext | Previous forecasting behaviours | Mean of 'TotalCost' over perious questions |
| **fprevavgEnhancedFrequency** | 124.540128 | Ext | Number/frequency of previous forecasts | Mean of 'EnhancedFrequency' over previous questions |
| **fprevavgRawFrequency** | 121.0447049 | Ext | Number/frequency of previous forecasts | Mean of 'RawFrequency' over previous questions |
| **MeanUpdateSize** | 73.08257418 | Int | Update frequency/size | Mean update size (set to a number outside the range of 0-1 if there have been no updates) |

[a] Internal (Int) variables refers to those which are generated within the given market under study. External (Ext) variables refer to those which are generated from markets external to the given market under study. These include variables from past markets and concurrent markets. The final full model is based on 43 features (19 internal features, 24 external features).

All available data were utilised and there was very little missing data.

**Next Generation Social Science pooled prediction markets -the external replication sample**

This pooled study consisted of four large-scale replication projects: the Reproducibility Project: Psychology;[8,9] the Experimental Economics Replication Project;[10] the Many Labs 2 project;[11,12] and the Social Science Replication Project.[13] In these projects, academics were asked to forecast the likelihood that a specific study would be able to be replicated. The pooled dataset had 103 markets (Supplementary Table 1). Across all markets, the absolute error of the prediction market was lower ($p<0.001$) than for an accompanying survey.[14] Participants were recruited via social media and mailing lists with the focus on people working within academia. The trading system was also an operator-staked LMSR platform with an inital token allocation and token gains converted to monetary rewards.[14] The LMSR liquidity factor was different in each of the four studies comprising the NGS2 dataset. Accordingly, in order to calculate point investment (and other similar metrics) in a consistent way, we used implied values (from the before and after trade probabilities) based on a liquidity factor of 150 to match the Almanis dataset. A key design difference to Almanis was that forecasters were expected to forecast based on their expertise and the information provided at market open (the publication containing the key finding and the planned study protocol for the replication study) on whether a study finding was expected to replicate or not. This meant there was a lesser reliance on new information emerging during the active market leading to reactive trading. Further the markets were of shorter duration with a mean of 13 (SD 1.2) days (Supplementary Table 1). Further details are available elsewhere.[8-10,12-14]

**Analysis**

**Forecaster accuracy model: development (Phase 1)**

**The relative Brier accuracy score as a measure of forecaster accuracy.**

For each trade, we subtract the Brier score of the market after the trade from the Brier score of the market before to obtain a relative Brier score.[3] We sum these trade-level relative Brier scores to generate a relative Brier accuracy probability for each forecaster on each question. The mean relative Brier was used rather than the median because, assessed by median, the Brier score ceases to be a proper scoring rule.[15]

**Past work rationale for variable selection**

In Phase 1 (forecaster accuracy model: development and validation), we trained a random forest model to identify in real-time forecasters likely to be either particularly accurate or particularly inaccurate based on variables (features) within the trading dataset. These variables were largely chosen by reviewing the literature on indicators of forecasting accuracy. We also incorporated some other measures less discussed in the literature which we believed may reflect user confidence in their own forecast. Atanasov et al.[16] showed that accurate forecasters tend to update their beliefs in frequent, small steps. We therefore included both forecasting frequency and mean update size as predictor variables. We also included the raw number of trades a user made on a question, as this may indicate engagement in the question and activity.[17] Atanasov et al.[16] further found that low-skill forecasters were more likely to reconfirm their initial forecasts, not updating them in the face of new information. This may indicate that these forecasters were more susceptible to confirmation bias[18] - the tendency to seek information that confirms initial beliefs. Therefore, we included a variable to measure the propensity of users to change their mind and forecast in the opposite direction to their initial forecast. Informed by the knowledge that open-minded forecasters tend to do better,[19] we considered that forecasters who were more confident in their forecasts may be more likely to make consecutive trades in the same direction and invest more points on their forecasts. Confident forecasters are likely to be more accurate than uncertain forecasters,[20] so we also included variables indicating how much a user spent as well as whether a user made a number of consecutive trades in the same direction on a question. We also considered features previously associated with insider trading.[21]

Many studies, such as those undertaken using the Good Judgement platform,[22] use weighted survey methods to create a crowd forecast. In these instances, users specify a probability distribution which serves as their forecast. However, as the Almanis platform uses a prediction market, users are essentially betting on a particular outcome when they make forecasts. Therefore, we can talk about the *direction* of their forecasts, which refers to whether they were betting on a yes or no outcome. This also gives us information on their expected probability of the outcome occurring, as the logarithmic market scoring rule used on Almanis incentives users to keep trading in a market until the market price matches their expectation.

**Development of the random forest model for grading forecast accuracy (Phase 1)**

The random forest approach creates an ensemble of different decision trees and then uses majority 'voting' of all the trees' outcomes to classify individual observations into discrete categories[23,24] with an embedded feature selection method which obtains very good performance (especially when considering out-of-sample performance), even in high-dimensional data when most of the features are 'noisy' (irrelevant to the outcome).[25,26] The random forest consisted of 500 decision trees (ntree = 500) while hyperparameter tuning occurred during cross-validation for the 'mtry' parameter (which determines how many variables are sampled at each split in each decision tree). The final value for mtry in the full model was 15. This trained the model on real-time data rather than simply focusing on summary data after a forecaster had made all their trades and also avoided the model sampling multiple performances from the same forecaster on the same question.

Evaluation of the original model on the training dataset after cross-validation was reported in Supplementary Fig. 1 legend. ROC curves[27] plot the yes positive rate (TPR), or sensitivity, against the no positive rate (FPR), which is equivalent to 1-specificity, for each different decision threshold for the outcome of being in the top quintile of the relative Brier accuracy score, above 0.024. This relative Brier cut-off was equivalent to moving the market probability by 1.2% or more when the pre-trade market probability was 50%. If forecasters did not move the crowd forecast very much with their trades, it was impossible for them to achieve a relative Brier score above 0.024 even if their forecasts were accurate. We have categorised these forecasters as "low volume traders" and they were excluded from Phase 1 (forecaster accuracy model: development).

**Evaluation of event prediction models taking into account forecaster accuracy rating (Phase 1)**

We then assessed how event prediction performance varied by forecaster accuracy rating using several methods. We graded forecasts in Almanis A and NGS2 by the machine accuracy rating of each selected trade. We developed four grades: Grade 1, scores 0.75 > 100; Grade 2, 0.5 > 0.75; Grade 3, 0.25 > 0.5; Grade 4, 0 > 0.25. High quality trades were those with a machine rating above

0.5. To assess differences in predictive effect by grade, we first examined additive effects using a generalised additive model. To assess multiplicative interaction, we added a product term for post trade probability and machine rated quality in a multivariable logistic model (Fig. 2C). This product term controlled for the effect that high quality forecasters may be likely to enter more difficult or uncertain markets. Instantaneous trades can provide information on the relative accuracy of the after trade compared to the prior trade.[28] Such AUC gain measures are independent of time to settlement. The latter is inversely related to forecast accuracy in our own and other[29] work. We compared the AUCs after and before trade (Fig. 3A), assessing the AUC gain (Fig. 3B) for trades stratified by grade. Further, hierarchical linear regression, clustering by market, was used for continuous measures such as the relative Brier accuracy probability. A similar model, clustered on the individual forecaster rather than market, was used to account for the potential contribution of inter-individual variation to accuracy.

**Development and optimisation of the weighted predictive model: The Hybrid human-machine event prediction model (Phase 2)**

We sought to build a model which would weight high quality forecasts more heavily and put less weight on lower machine quality rated 'Grade 4' forecasters, who generally made market prices less predictive of event occurrence.

Our method of reweighting forecasts relied on the structure of the logarithmic market scoring rule (LMSR) for prediction markets. In such markets, the price of a 'yes' contract (a contract which pays out 1 point if an event occurs and nothing if not) reflects the aggregated expectation that an event will occur. This price is calculated according to the following equation in LMSR markets:

$$P = e^{N_T/B}/(e^{N_T/B} + e^{N_F/B}) \qquad (1)$$

where P is the market price of yes contracts, $N_T$ and $N_F$ are, respectively, the number of yes and the number of no contracts that have been purchased and B is a liquidity factor (set to 150 for all questions on the Almanis platform).

Our method to provide more accurate probabilities involved weighting yes and no contracts from high quality forecasters more heavily, and those from Grade 4 forecasters less heavily to calculate an enhanced market price P'. However, it was not clear how much more or for how long these forecasts should be weighted differently to achieve the best results.

Therefore, we tested a number of different models using five-fold cross validation repeated three times on the Almanis A dataset. Each model was assessed on the improvement in the mean market Brier score (calculated across all markets on the platform) and the model which improved the mean market Brier score the most was chosen as the final model. We then validated the final model on the Almanis B and NGS2 datasets.

Creating the final model involved optimising over three steps:

1. Training a model which weighted high quality forecasters' contracts more heavily to generate more accurate market probabilities
2. Training a model which weighted Grade 4 forecasters' contracts less heavily to generate more accurate market probabilities
3. Combining the models from steps 1 and 2 and recalibrating the results to account for any distortionary effects of adding or removing contracts form the market.

In the first step, multiple models were assessed. The model parameters were optimised using a grid search and the cross validation assessed the performance of the model. If the grid search indicated best performance at the edge of the parameter space, the cross validation was repeated with a wider parameter space in an attempt to ensure that the best possible model was chosen. The four major model types are laid out below.

Model A – This model weighted high quality forecasters' contracts more heavily in a linear fashion. Let $\mathrm{HI}_c^{\mathrm{i,T}}$ and $\mathrm{HI}_c^{\mathrm{i,F}}$ be, respectively, the number of yes and no contracts bought by the $i^{th}$ high-quality trader in the last C trades (across the market) and let

$$\mathrm{HI}_c^i = \mathrm{HI}_c^{\mathrm{i,T}} - \mathrm{HI}_c^{\mathrm{i,F}}$$

be the net number of yes contracts bought by the $i^{th}$ high-quality trader. Then the enhanced market probability P' was calculated by the following adaption of equation 1:

$$P' = \frac{e^{(N_T + x * \sum_i HI_\infty^i)/B}}{(e^{(N_T + x * \sum_i HI_\infty^i)/B} + e^{N_F/B})} \quad (2)$$

This made contracts purchased by high quality forecasters' worth (1+x) times as much as other contracts when calculating enhanced market probabilities.

Model B - This model weighted high quality forecasters' contracts linearly (like Model A), but with a parameter capping how many trades this weighting was carried forward for.

Model C – This model was an adaption of Model B with an additional a parameter capping how much more heavily an individual trade could be weighted.

Model D – This model was an adaption of Model C with an additional parameter capping the total number of weighted contracts across the market.

We also trialled each of these models with a further parameter allowing the exponential decay of additional weighting over future trades.

Similar models were used in step 2. Instead of weighting contracts bought by high quality forecasters more highly, we underweighted contracts bought by Grade 4 forecasters, for instance by considering negative values of *x* in an analogue of equation 2.

Model C with no exponential decay was the best performing model in cross validation step 1 and an analogue of Model B with exponential decay was the best performing Model in cross validation in step 2. The best parameters for these models across all of the Almanis A dataset were then found using grid search.

These models were then combined in step 3. Contracts were weighted more heavily when bought by high quality forecasters *and* contracts bought by Grade 4 forecasters were weighted less heavily to create a single enhanced market probability P''. Ideally steps 1 and 2 would be completed together, but this would require a prohibitive amount of computational power due to the high number of parameters. Therefore, there was a final optimisation step to reduce any tendency for probabilities to become extreme by *both* overweighting some contracts and underweighting others.

The final probabilities in step 3 were of the form:

$$P^E = \begin{cases} P & if\ |P - P''| \leq h \\ 0.5 + g * (P'' - 0.5) & if\ |P - P''| > h \end{cases} \quad (3)$$

where $P^E$ is the final enhanced probability, P is the market probability, P'' is as defined above and *g* and *h* were parameters to be optimised using grid search. If the market and enhanced probabilities were sufficiently close, the final enhanced model reverted to the market probability, and if the difference was greater, probabilities given by the enhanced model were de-extremised.

Each of steps 1, 2 and 3 were shown to improve predictive performance in cross validation, which justified their inclusion in the final model. In the final enhanced prediction model, contracts bought by high quality forecasters were weighted at 1.425 times their usual weight until 20 further trades had been made in the market, provided less than 55 contracts were bought in a single trade. If more than 55 were bought in a single trade, these contracts were weighted normally. Grade 3 and low volume trades with scores of 0.5 or less were not weighted. Contracts bought by Grade 4 forecasters were weighted at 0.285 times their usual weight initially. This 71.5% reduction in weighting decayed exponentially at a rate of 12% per trade until vanishing after 16 trades. In step 3, the final parameter choices for equation 3 after grid search across the Almanis A dataset were h = 0.0235 and g = 0.8575. This final enhanced model was then validated on the Almanis B and NGS2datasets using these values optimised on Almanis A.

Grade 1 trades were overweighted 1.425- fold until 20 further trades had been made. To reduce the likelihood that the market would be pushed towards overly extreme probabilities, the small number of trades with 55 or more points remained weighted normally. Grade 3 and low volume trades with scores of 0.5 or less were not weighted. Grade 4 trades weighted 0.285- fold with a forward exponential decay of 12% per trade for 16 further trades.

**COVID-19 Event Prediction**

We used a hierarchical regression model to allow intra-market comparisons of the hybrid vs. general trend for mean absolute prediction error against time to settlement.[30]

**Table S1. Characteristics of the key prediction market samples.**

| | Almanis Training | Almanis Recent A | Almanis Recent B | NGS2 Pooled |
|---|---|---|---|---|
| **Date of commencement** | 09/11/2017 | 01/03/2020 | 01/01/2021 | 19/11/2012 |
| **Date of cessation** | 29/02/2020 | 31/12/2020 | 30/06/2021 | 22/11/2016 |
| **Market number** | 766 | 614 | 282 | 103 |
| **Market duration (mean in days (SD))** | 64 (70) | 55 (52) | 39.8 (26.3) | 13 (1.2) |
| **Forecaster number** | 584 | 614 | 506 | 347 |
| **Trade number** | 60,857 | 52,173 | 19,933 | 7,849 |
| **Trader per market (Median [iqr] (full))** | 42 [31-55] (8-142) | 37 [25-54] (3-176) | 27 [18-43] (2-121) | 37 [27-46] (18-68) |
| **Trades per market (Median [iqr] (full))** | | 62 [38-105.8] (3-917) | 45 [26-81.5] (2-408) | 70 [52-90] (26-193) |
| **Markets per trader (Median [iqr] (full))** | 22 [6-65] (1-739) | 19 [6-48] (1-527) | 8 [3-19] (1-245) | 10 [6-15] (1-35) |
| **Trades per trader (Median [iqr] (full))** | 26 [7-76] (1-6192) | 26 [9-61] (1-4306) | 13 [6-33] (1-2560) | 13 [8-24] (1-311) |
| **Spearman correlation of likelihood to settlement (95% CI)** | 0.604 (0.599, 0.609) | 0.572 (0.566, 0.578) | 0.54 (0.53, 0.55) | 0.57 (0.55, 0.58) |
| **General AUC (95% CI)** | 0.918 (0.915, 0.92) | 0.891 (0.887, 0.894) | 0.915 (0.910, 0.921) | 0.827 (0.818, 0.836) |
| **Relative Brier Score (mean (SD))** | 0.005 (0.13) | 0.0022 (0.13) | 0.0018 (0.12) | 0.0015 (0.08) |
| **% Extreme initial probability (<20,>80%)** | 66.8% (40627/60857) | 73.1% (38117/52173) | 75.7% (15099/19933) | 0% (0/71849) |

Note: COVID-19 topic markets not shown. Overall Almanis market number = 1822.

**Table S2. Characteristics of the prediction market samples by trade probability set at market start.**

| | Almanis A | | Almanis B | | NGS2 |
|---|---|---|---|---|---|
| | Intermediate baseline probability 20%-80% | Extreme baseline probability <20%, >80% | Intermediate baseline probability 20%-80% | Extreme baseline probability <20%, >80% | Intermediate baseline probability 20%-80%* |
| **Market number** | 116 | 498 | 41 | 241 | 103 |
| **Forecaster number** | 506 | 597 | 322 | 491 | 347 |
| **Trade number** | 12303 | 39870 | 4211 | 15722 | 7849 |
| **Overall** | | | | | |
| **AUC (95%CI)** | 0.853 (0.781, 0.924) | 0.892 (0.841, 0.943) | 0.787 (0.637, 0.938) | 0.945 (0.910, 0.980) | 0.827 (0.763, 0.891) |
| **Mean Raw Brier (SD)** | 0.308 (0.437) | 0.182 (0.409) | 0.003 (0.137) | 0.001 (0.117) | 0.396 (0.361) |
| **Mean relative Brier (SD)** | 0.004 (0.155) | 0.002 (0.120) | 0.342 (0.482) | 0.107 (0.286) | 0.002 (0.080) |
| **Machine quality rating >0.5** | | | | | |
| **AUC (95%CI)** | 0.861 (0.791, 0.932) | 0.893 (0.846, 0.939) | 0.784 (0.623, 0.945) | 0.958 (0.927, 0.988) | 0.826 (0.758, 0.893) |
| **Mean Raw Brier (SD)** | 0.230 (0.436) | 0.199 (0.427) | 0.328 (0.487) | 0.089 (0.272) | 0.393 (0.366) |
| **Mean relative Brier (SD)** | 0.007 (0.148) | 0.006 (0.118) | 0.002 (0.128) | 0.020 (0.100) | 0.003 (0.095) |
| **Machine quality rating ≤0.5** | | | | | |
| **AUC (95%CI)** | 0.833 (0.755, 0.910) | 0.883 (0.814, 0.952) | 0.792 (0.653, 0.931) | 0.922 (0.874, 0.970) | 0.828 (0.760, 0.896) |
| **Mean Raw Brier (SD)** | 0.328 (0.438) | 0.155 (0.378) | 0.316 (0.472) | 0.130 (0.301) | 0.399 (0.358) |
| **Mean relative Brier (SD)** | -0.003 (0.169) | -0.004 (0.122) | 0.006 (0.152) | -0.023 (0.133) | 0.0002 (0.067) |
| **Hybrid** | | | | | |
| **AUC (95%CI)** | - | - | 0.793 (0.640, 0.947) | 0.958 (0.925, 0.992) | 0.834 (0.771, 0.897) |

*NGS2 has no questions with extreme initial baseline probability <20%, >80%).

**Table S3. A reduction in time to settlement is associated with improved market accuracy.**

| Time of settlement (days) | % of total trades | No. of trades | AUC (After trade) * | Raw Brier Mean (SD) | Relative Brier Mean (SD) |
|---|---|---|---|---|---|
| **Almanis-A** | | | | | |
| 0 – 2 | 13.28 | 6929 | 0.951 (0.912, 0.989) | 0.16 (0.36) | 0.0013 (0.14) |
| 3 – 6 | 10.68 | 5570 | 0.946 (0.909, 0.983) | 0.16 (0.32) | 0.0021 (0.15) |
| 7 – 14 | 19.47 | 10158 | 0.887 (0.842, 0.933) | 0.24 (0.45) | 0.0029 (0.15) |
| 15 – 29 | 27.21 | 14194 | 0.859 (0.795, 0.923) | 0.22 (0.45) | 0.0013 (0.12) |
| 30 – 59 | 23.11 | 12057 | 0.858 (0.793, 0.923) | 0.19 (0.41) | 0.0033 (0.10) |
| ≥60 | 6.26 | 3265 | 0.773 (0.684, 0.862) | 0.35 (0.43) | 0.0016 (0.15) |
| *P* trend over decreasing category** | | | | **$<2 \times 10^{-16}$** | 0.63 |
| **Almanis-B** | | | | | |
| 0 – 2 | 19.5 | 3886 | 0.951 (0.893, 1.000) | 0.09 (0.23) | 0.001 (0.11) |
| 3 – 6 | 14.8 | 2948 | 0.938 (0.893, 984) | 0.14 (0.32) | 0.001 (0.13) |
| 7 – 14 | 24.6 | 4894 | 0.889 (0.835, 0.944) | 0.20 (0.40) | 0.003 (0.15) |
| 15 – 29 | 22.6 | 4512 | 0.887 (0.808, 0.966) | 0.18 (0.41) | 0.0001 (0.12) |
| 30 – 59 | 16.1 | 3217 | 0.927 (0.864, 0.989) | 0.11 (0.29) | 0.003 (0.09) |
| ≥60 | 2.4 | 476 | 0.921 (0.832, 1.011) | 0.21 (0.38) | 0.01 (0.10) |
| *P* trend over decreasing category** | | | | **$2.40 \times 10^{-4}$** | 0.12 |
| **NGS2** | | | | | |
| 0 – 2 | 16.80 | 1319 | 0.823 (0.734, 0.912) | 0.40 (0.42) | -0.0003 (0.05) |
| 3 – 6 | 21.70 | 1703 | 0.841 (0.761, 0.922) | 0.38 (0.37) | 0.0006 (0.07) |
| 7 – 14 | 61.50 | 4827 | 0.822 (0.756, 0.887) | 0.40 (0.34) | 0.002 (0.09) |
| 15 – 29 | - | - | - | - | - |
| 30 – 59 | - | - | - | - | - |
| ≥60 | - | - | - | - | - |
| *P* trend over decreasing category** | | | | 0.94 | 0.08 |

Note 1: The NGS platform had earlier trading compared to the Almanis platform with 43.0% of trades occurring in the first three days vs 29.9% in Almanis B.
Note 2: The relative Brier accuracy probability was independent of this time-to-settlement accuracy improvement and thus preferentially used where time-to-settlement effects on predictive performance needed to be controlled for.
Note 3: This is consistent with past work[29].

**Table S4. Data table underlying Fig. S2. The predictive performance for event occurrence is higher with increasing machine rating of trade quality in Almanis A and NGS2.**

| Grades (informativeness score range) | Number of trades % (n/N) | AUC after trade (95% CI) | AUC before trade (95% CI) | AUC temporal gain % (95% CI)^ | p Value* | Relative Brier accuracy score Mean (SD) |
|---|---|---|---|---|---|---|
| **Almanis A** | | | | | | |
| Grade 1 (0.75-1)[a] | 16.3% (8488/52173) | 0.865 (0.813, 0.914) | 0.857 (0.807, 0.907) | 0.70 (0.3, 1.00) | **0.001** | 0.009 (0.09) |
| Grade 2 (0.5-0.75)[a] | 20.5% (10712/52173) | 0.789 (0.722, 0.858) | 0.777 (0.711, 0.843) | 1.30 (0.80, 1.80) | **$3.64 \times 10^{-7}$** | 0.011 (0.19) |
| Grade 3 (0.25-0.5)[b] | 8.8% (4582/52173) | 0.746 (0.698, 0.794) | 0.744 (0.694, 0.795) | 0.20 (-0.80, 1.10) | 0.77 | 0.007 (0.22) |
| Grade 4 (0-0.25)[b] | 6.2% (3234/52173) | 0.829 (0.786, 0.872) | 0.868 (0.829, 0.907) | -3.90 (-5.30, -2.60) | **$6.15 \times 10^{-9}$** | -0.035 (0.19) |
| Low volume, machine quality rating >0.5[c] | 25.1% (13093/52173) | 0.926 (0.883, 0.970) | 0.926 (0.882, 0.970) | 0.01 (-0.10, 0.20) | 0.879 | 0.0002 (0.05) |
| Low volume, machine quality rating ≤0.5[d] | 23.1% (12064/52173) | 0.880 (0.779, 0.981) | 0.879 (0.779, 0.980) | 0.03 (-0.20, 0.20) | 0.79 | $-3.06 \times 10^{-5}$ (0.02) |
| Machine quality rating >0.5[a+c] | 61.9% (32293/52173) | 0.889 (0.855, 0.932) | 0.884 (0.849, 0.927) | 0.60 (0.40, 0.80) | **$9.30 \times 10^{-9}$** | 0.006 (0.13) |
| Machine quality rating ≤0.5[b+d] | 38.1% (19880/52173) | 0.888 (0.837, 0.939) | 0.891 (0.844, 0.924) | -0.30 (-0.60, -0.06) | **0.002** | -0.005 (0.13) |
| All trades | 52173 | 0.891 (0.850, 0.932) | 0.888 (0.847, 0.929) | 0.30 (0.20, 0.30) | **$9.13 \times 10^{-19}$** | 0.002 (0.13) |
| **NGS2** | | | | | | |
| Grade 1 (0.75-1) | 0.45% (35/7849) | 0.563 (0.328, 0.798) | 0.546 (0.369, 0.723)* | 1.72 (-13.3, 16.75) | 0.82 | 0.051 (0.14) |
| Grade 2 (0.5-0.75) | 18.6% (1480/7849) | 0.764 (0.671, 0.857) | 0.751 (0.656, 0.845) | 1.34 (0.41, 2.28) | **0.005** | 0.004 (0.12) |
| Grade 3 (0.25-0.5) | 32.8% (2573/7849) | 0.781 (0.701, 0.862) | 0.777 (0.697, 0.856) | 0.46 (-0.10, 1.03) | 0.11 | 0.002 (0.08) |
| Grade 4 (0 – 0.25) | 1.6% (124/7849) | 0.814 (0.659, 0.968) | 0.864 (0.732, 0.99) | -5.04 (-1.59, 11.70) | 0.14 | -0.034 (0.15) |
| Low volume, machine quality rating >0.5[c] | 22.9% (1800/7849) | 0.869 (0.801, 0.937) | 0.867 (0.798, 0.936) | 0.20 (-0.18, 0.58) | 0.30 | 0.0017 (0.06) |
| Low volume, machine quality rating ≤0.5[d] | 23.4% (1837/7849) | 0.867 (0.797, 0.936) | 0.867 (0.798, 0.937) | 0.007 (-0.10 , 0.25) | 0.42 | $-3.08 \times 10^{-5}$ (0.03) |
| Machine quality rating >0.5[a+c] | 42.2% (3315/7849) | 0.826 (0.758, 0.893) | 0.820 (0.751, 0.888) | 0.62 (0.19, 1.05) | **0.005** | $3.29 \times 10^{-3}$ (0.095) |
| Machine quality rating ≤0.5[b+d] | 57.8% (4534/7849) | 0.828 (0.760, 0.896) | 0.826 (0.759, 0.894) | 0.13 (-0.23, 0.48) | 0.479 | $3.29 \times 10^{-3}$ (0.067) |

| | | | | | | |
|---|---|---|---|---|---|---|
| All trades | 7849 | 0.827 (0.763, 0.891) | 0.824 (0.759, 0.888) | 0.34 (0.19, 0.49) | **$7.47x10^{-6}$** | 0.0015 (0.08) |

**Footnote:** Low volume traders were those whose forecasts did not move the market very much. The cut-off for this classification was a potential maximum relative Brier score of 0.024 on a question, which was equivalent to moving the market forecast by 1.2% when the market probability was 50%[a]. Grades 1 and 2, [b] refers to grades 3 and 4, [c] refers to low volume with machine quality rating >0.5, [d] low volume and machine quality rating <=0.5. *p values for difference in after vs. before AUC, clustered within markets. Machine rating refers to machine rating of trade quality. For low volume with a machine rating of 0.5 or less, we removed them from a 'high quality' binary classification because a low machine quality rating may have been due to the low volume. Therefore, 'high quality' in the paper refers to a+c =1 vs. b=0. All clustered within markets.* Grade 1 trades are being preferentially made in uncertain difficult markets, that is, markets with a lower AUC before trade. ^% AUG temporal gain = gain×100.

**Table S5. The evolution of higher trading accuracy as time to settlement is anticipated earlier by high quality trades in Almanis A.**

| Days to settlement | Trade categories for human forecasts by machine rating | Number of forecasts | Percentage of forecasts of that category (%) | Relative Brier accuracy score (Mean (SD)) | % AUC gain (95% CI) | p Value |
|---|---|---|---|---|---|---|
| 0-2 | Human only forecast | 6929 | 100% | 0.001 (0.144) | 0.11 (-0.01, 0.22) | 0.074 |
| | Machine quality rating >0.5 | 5320 | 76.8% | 0.006 (0.168) | 0.45 (-0.19, 1.09) | 0.173 |
| | Machine quality rating ≤0.5 | 1609 | 23.2% | -0.014 (0.135) | 0.91 (-0.20, 2.02) | 0.109 |
| 3-6 | Human only forecast | 5570 | 100% | 0.002 (0.145) | 0.15 (0.03, 0.027) | **0.014** |
| | Machine quality rating >0.5 | 4108 | 73.8% | 0.008 (0.137) | 0.34 (0.04, 0.65) | **0.026** |
| | Machine quality rating ≤0.5 | 1462 | 26.2% | -0.013 (0.165) | -0.80 (-0.04, 1.64) | 0.061 |
| 7-14 | Human only forecast | 10158 | 100% | 0.003 (0.147) | 0.27 (0.18, 0.35) | **$7.312 \times 10^{-10}$** |
| | Machine quality rating >0.5 | 7398 | 72.8% | 0.003 (0.134) | 0.43 (0.22, 0.63) | **$3.711 \times 10^{-5}$** |
| | Machine quality rating ≤0.5 | 2760 | 27.2% | 0.003 (0.179) | -0.38 (-0.24, 0.10) | 0.233 |
| 15-29 | Human only forecast | 14194 | 100% | 0.001 (0.117) | 0.17 (0.09, 0.26) | **$9.05 \times 10^{-5}$** |
| | Machine quality rating >0.5 | 7760 | 54.7% | 0.006 (0.116) | 0.43 (0.12, 0.75) | **0.007** |
| | Machine quality rating ≤0.5 | 6434 | 45.3% | -0.005 (0.118) | -0.15 (-0.5, 0.16) | 0.337 |
| 30-59 | Human only forecast | 12057 | 100% | 0.003 (0.100) | 0.56 (0.34, 0.78) | **$6.325 \times 10^{-7}$** |
| | Machine quality rating >0.5 | 5624 | 46.6% | 0.008 (0.107) | 0.94 (0.43, 1.45) | **0.0003** |
| | Machine quality rating ≤0.5 | 6433 | 53.4% | -0.001 (0.094) | 0.39 (-0.38, 1.15) | 0.321 |
| 60+ | Human only forecast | 3265 | 100% | 0.002 (0.149) | 0.20 (-0.15, 0.56) | 0.258 |
| | Machine quality rating >0.5 | 2083 | 63.8% | 0.009 (0.145) | 0.24 (-0.50, 0.98) | 0.524 |
| | Machine quality rating ≤0.5 | 1182 | 36.2% | -0.012 (0.156) | -0.79 (-1.87, 0.30) | 0.152 |

Note: As markets become more accurate towards settlement, the high accuracy quality trades provide a time advantage. At day 20, these markets provide similar accuracy metrics as for other trades 0-2 days from market settlement.

**Table S6. Data table underlying Fig. 3. Performance of the hybrid vs. human only models stratified by disagreement level.**

| Disagreement Level at the time of trade | | AUC (95% CI) | | | | Mean absolute prediction error (Std error) | | |
|---|---|---|---|---|---|---|---|---|
| | | Human only | Hybrid | % AUC gain for Hybrid vs. human only after trade | p Value for difference | Human only | Hybrid | p Value for difference |
| **Almanis B** | | | | | | | | |
| **≤ 5%** | **80.9% (16129/19933)** | 0.940 (0.902, 0.977) | 0.940 (0.903, 0.978) | 0.03 (-0.07, 0.14) | 0.509 | 0.0906 (0.0095) | 0.093 (0.0094) | **0.013** |
| **More than 5%** | **19.1% (3804/19933)** | 0.768 (0.716, 0.820) | 0.904 (0.853, 0.948) | 13.21 (9.85, 16.58) | **$1.35x10^{-14}$** | 0.343 (0.0083) | 0.216 (0.0121) | **$<2.00x10^{-16}$** |
| **More than 10%** | **11.8% (2351/19933)** | 0.707 (0.655, 0.759) | 0.913 (0.866, 0.960) | 20.57 (16.42, 24.72) | **$<1.00x10^{-15}$** | 0.388 (0.0083) | 0.223 (0.0139) | **$<2.00x10^{-16}$** |
| **More than 15%** | **7.6% (1517/19933)** | 0.629 (0.570, 0.687) | 0.930 (0.875, 0.984) | 30.09 (23.60, 36.60) | **$<1.00x10^{-15}$** | 0.422 (0.0076) | 0.215 (0.0136) | **$<2.00x10^{-16}$** |
| **More than 20%** | **5.1% (1009/19933)** | 0.502 (0.428, 0.576) | 0.950 (0.893, 1.000) | 44.76 (37.61, 51.90) | **$<1.00x10^{-15}$** | 0.462 (0.0091) | 0.204 (0.0135) | **$<2.00x10^{-16}$** |
| **More than 30%** | **1.9% (398/19933)** | 0.137 (0.107, 0.167) | 0.969 (0.920, 1.000) | 83.26 (76.83, 89.69) | **$<1.00x10^{-15}$** | 0.558 (0.0090) | 0.200 (0.0136) | **$<2.00x10^{-16}$** |
| **More than 40%** | **0.6% (122/19933)** | 0.039 (-0.008, 0.086) | 0.964 (0.894, 1.000) | 92.44 (81.04, 100) | **$<1.00x10^{-15}$** | 0.625 (0.0183) | 0.210 (0.0259) | **$<2.00x10^{-16}$** |
| **All trades** | | 0.915 (0.873, 0.958) | 0.931 (0.891, 0.972) | 1.60 (1.06, 2.14) | **$6.104x10^{-9}$** | 0.114 (0.0092) | 0.104 (0.0093) | **$<2.00x10^{-16}$** |
| **NGS2** | | | | | | | | |
| **≤ 5%** | **92.0% (7222/7849)** | 0.837 (0.775, 0.899) | 0.837 (0.775, 0.899) | 0.01 (-0.27, 0.28) | 0.967 | 0.403 (0.018) | 0.401 (0.018) | 0.332 |
| **More than 5%** | **7.99% (627/7849)** | 0.601 (0.408, 0.794) | 0.739 (0.583, 0.895) | 13.80 (4.61, 23.0) | **0.003** | 0.454 (0.027) | 0.432 (0.024) | **$5.52x10^{-15}$** |
| **More than 10%** | **0.64% (50/7849)** | 0.478 (0.085, 0.870) | 0.800 (0.346, 1.000) | 32.22 (11.28, 53.17) | **0.003** | 0.501 (0.055) | 0.433 (0.038) | **$1.17x10^{-8}$** |
| **All trades** | | 0.827 (0.763, 0.891) | 0.833 (0.767, 0.896) | 0.71 (-0.03, 1.38) | **0.039** | 0.405 (0.018) | 0.402 (0.018) | **0.0098** |

**Table S7. Data table underlying Fig. 3. Performance of the hybrid vs. human only models stratified by disagreement level.**

| **Disagreement level at the time of trade** | | **AUC (95% CI)** | | | | **Mean absolute prediction error (Std error)** | | |
|---|---|---|---|---|---|---|---|---|
| | | **Human only** | **Hybrid** | **% AUC gain for hybrid vs. human only after trade** | **p Value for difference** | **Human only** | **Hybrid** | **p Value for difference** |
| **Almanis B** | | | | | | | | |
| ≤ 5% | 80.9% (16129/19933) | 0.940 (0.902, 0.977) | 0.940 (0.903, 0.978) | 0.03 (-0.07, 0.14) | 0.509 | 0.0906 (0.0095) | 0.093 (0.0094) | **0.013** |
| More than 5% | 19.1% (3804/19933) | 0.768 (0.716, 0.820) | 0.904 (0.853, 0.948) | 13.21 (9.85, 16.58) | **1.35x10$^{-14}$** | 0.343 (0.0083) | 0.216 (0.0121) | **<2.00x10$^{-16}$** |
| More than 10% | 11.8% (2351/19933) | 0.707 (0.655, 0.759) | 0.913 (0.866, 0.960) | 20.57 (16.42, 24.72) | **<1.00x10$^{-15}$** | 0.388 (0.0083) | 0.223 (0.0139) | **<2.00x10$^{-16}$** |
| More than 15% | 7.6% (1517/19933) | 0.629 (0.570, 0.687) | 0.930 (0.875, 0.984) | 30.09 (23.60, 36.60) | **<1.00x10$^{-15}$** | 0.422 (0.0076) | 0.215 (0.0136) | **<2.00x10$^{-16}$** |
| More than 20% | 5.1% (1009/19933) | 0.502 (0.428, 0.576) | 0.950 (0.893, 1.000) | 44.76 (37.61, 51.90) | **<1.00x10$^{-15}$** | 0.462 (0.0091) | 0.204 (0.0135) | **<2.00x10$^{-16}$** |
| More than 30% | 1.9% (398/19933) | 0.137 (0.107, 0.167) | 0.969 (0.920, 1.000) | 83.26 (76.83, 89.69) | **<1.00x10$^{-15}$** | 0.558 (0.0090) | 0.200 (0.0136) | **<2.00x10$^{-16}$** |
| More than 40% | 0.6% (122/19933) | 0.039 (-0.008, 0.086) | 0.964 (0.894, 1.000) | 92.44 (81.04, 100) | **<1.00x10$^{-15}$** | 0.625 (0.0183) | 0.210 (0.0259) | **<2.00x10$^{-16}$** |
| All trades | | 0.915 (0.873, 0.958) | 0.931 (0.891, 0.972) | 1.60 (1.06, 2.14) | **6.104x10$^{-9}$** | 0.114 (0.0092) | 0.104 (0.0093) | **<2.00x10$^{-16}$** |
| **NGS2** | | | | | | | | |
| ≤ 5% | 92.0% (7222/7849) | 0.837 (0.775, 0.899) | 0.837 (0.775, 0.899) | 0.01 (-0.27, 0.28) | 0.967 | 0.403 (0.018) | 0.401 (0.018) | 0.332 |
| More than 5% | 7.99% (627/7849) | 0.601 (0.408, 0.794) | 0.739 (0.583, 0.895) | 13.80 (4.61, 23.0) | **0.003** | 0.454 (0.027) | 0.432 (0.024) | **5.52x10$^{-15}$** |
| More than 10% | 0.64% (50/7849) | 0.478 (0.085, 0.870) | 0.800 (0.346, 1.000) | 32.22 (11.28, 53.17) | **0.003** | 0.501 (0.055) | 0.433 (0.038) | **1.17x10$^{-8}$** |
| All trades | | 0.827 (0.763, 0.891) | 0.833 (0.767, 0.896) | 0.71 (-0.03, 1.38) | **0.039** | 0.405 (0.018) | 0.402 (0.018) | **0.0098** |

**Table S8. Proportion of discordant (opposing) event calls between market and insight, observed and expected.**

| Absolute Difference | Proportion Observed[a] (n/N Trades) | Proportion Expected[b] (n/N Trades) | Proportion Observed[c] (n/N Markets) | Proportion Expected[d] (n/N Markets) | P-value (difference between observed and expected markets) |
|---|---|---|---|---|---|
| **Almanis B** | | | | | |
| Overall | 444/528 (84.1%) | 264/528 (50%) | 65/84 (77.4%) | 42/84 (50%) | $2.4 \times 10^{-7}$ |
| Less than 10% | 71/108 (65.7%) | 54/108 (50%) | 18/29 (62.1%) | 15/29 (50%) | 0.13 |
| 10% or more | 373/420 (88.8%) | 210/420 (50%) | 63/77 (81.8%) | 39/77 (50%) | $7.1 \times 10^{-9}$ |
| 20% or more | 325/344 (94.5%) | 177/344 (50%) | 56/60 (93.3%) | 30/60 (50%) | $4.5 \times 10^{-13}$ |
| 30% or more | 252/261 (96.6%) | 131/261 (50%) | 50/52 (96.2%) | 26/52 (50%) | $3.1 \times 10^{-13}$ |
| **Covid-19** | | | | | |
| Overall | 117/171 (68.4%) | 86/171 (50%) | 24/33 (72.7%) | 17/33 (50%) | 0.0068 |
| Less than 10% | 23/57 (40.4%) | 29/57 (50%) | 10/18 (55.6%) | 9/18 (50%) | 0.41 |
| 10% or more | 94/114 (82.5%) | 67/114 (50%) | 18/22 (81.8%) | 11/22 (50%) | 0.0022 |
| 20% or more | 61/64 (95.3%) | 32/64 (50%) | 14/15 (93.3%) | 8/15 (50%) | 0.00049 |
| 30% or more | 54/54 (100%) | 27/54 (50%) | 13/13 (100%) | 7/13 (50%) | 0.00012 |

[a] The proportion of opposing event calls where the insight is true, and the market is not true.
[b] The proportion of opposing event calls expected by chance where the insight is true, and the market is not true.
[c] The proportion of opposing event calls where the insight is true, and the market is not true, grouped by market.
[d] The proportion of opposing event calls expected by chance where the insight is true, and the market is not true, grouped by market.
Note: NGS2 data too sparse for reporting (see Table S7)

***Fig. S1. Predicting informative trades in the top quintile of relative Brier accuracy values in Almanis-Training and the Almanis A and NGS2 test prediction markets.***

**Panel A: Area under the curve for within-market, external, and full models in Almanis T, A, and NGS2.**

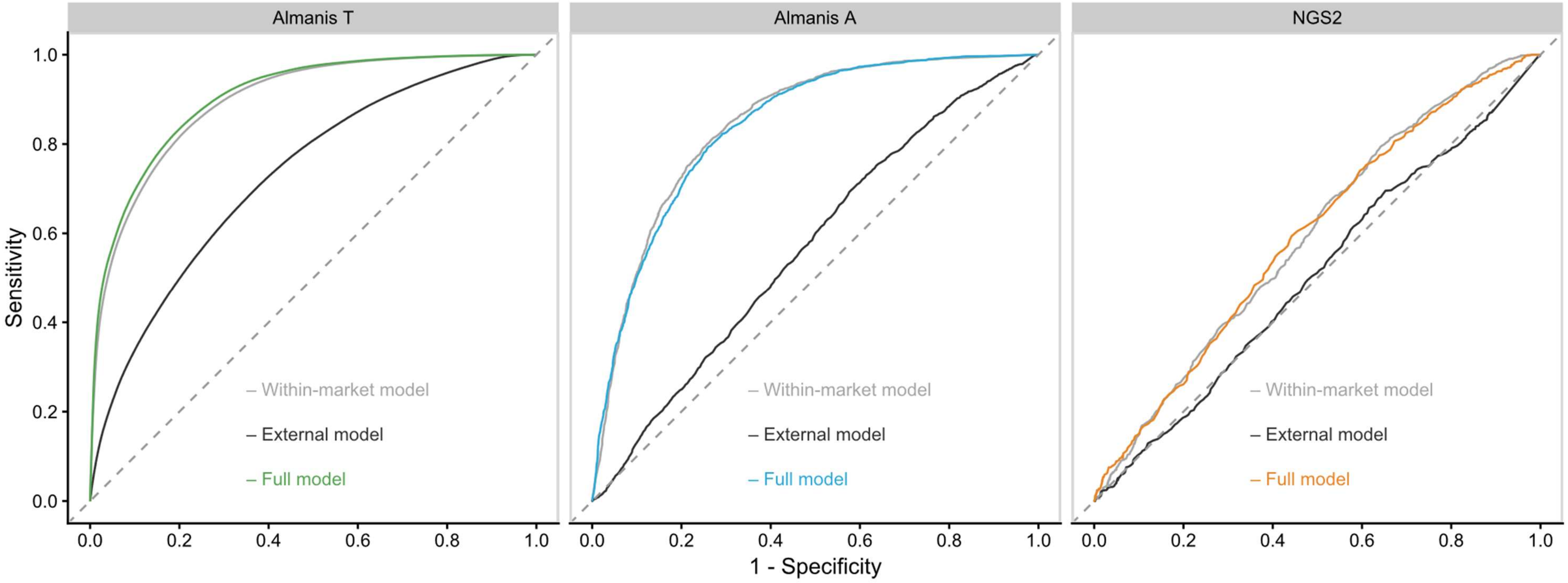

| Panel B: Accuracy performance metrics for within-market, external, and full models in Almanis T, A, and NGS2. | | | | | | | |
|---|---|---|---|---|---|---|---|
| **Model** | **Accuracy (95% CI)** | **Sensitivity (95% CI)** | **Specificity (95% CI)** | **PPV (95% CI)** | **NPV (95% CI)** | **AUC (95% CI)** | **p Value** |
| **Almanis training** | | | | | | | |
| **M1** (External to market) | 0.668 (0.659, 0.676) | 0.690 (0.679, 0.701) | 0.641 (0.629, 0.653) | 0.693 (0.682, 0.703) | 0.639 (0.626, 0.651) | 0.728 (0.720, 0.737) | **$<2x10^{-16}$** |
| **M2** (Within-market behaviour) | 0.817 (0.810, 0.824) | 0.851 (0.843, 0.860) | 0.777 (0.766, 0.788) | 0.817 (0.808, 0.826) | 0.817 (0.807, 0.827) | 0.899 (0.894, 0.904) | **$<2x10^{-16}$** |
| **All** (Combined **M1** + **M2**) | 0.822 (0.815, 0.828) | 0.855 (0.847, 0.864) | 0.782 (0.772, 0.793) | 0.822 (0.813, 0.830) | 0.822 (0.812, 0.832) | 0.906 (0.901, 0.911) | **$<2x10^{-16}$** |
| **Almanis A** | | | | | | | |
| **M1** (External to market) | 0.563 (0.552, 0.574) | 0.545 (0.534, 0.556) | 0.584 (0.573, 0.594) | 0.598 (0.588, 0.609) | 0.529 (0.518, 0.540) | 0.579 (0.566, 0.591) | **$<2x10^{-16}$** |
| **M2** (Within-market behaviour) | 0.775 (0.765, 0.784) | 0.855 (0.847, 0.863) | 0.683 (0.673, 0.693) | 0.755 (0.745, 0.764) | 0.805 (0.796, 0.814) | 0.845 (0.836, 0.853) | **$<2x10^{-16}$** |
| **All** (Combined **M1** + **M2**) | 0.769 (0.759, 0.778) | 0.826 (0.818, 0.835) | 0.703 (0.693, 0.713) | 0.761 (0.751, 0.770) | 0.781 (0.771, 0.790) | 0.840 (0.831, 0.848) | **$<2x10^{-16}$** |
| **Test- NGS2** | | | | | | | |
| **M1** (External to market) | 0.580 (0.559, 0.602) | 0.668 (0.648, 0.688) | 0.477 (0.456, 0.499) | 0.599 (0.578, 0.620) | 0.552 (0.531, 0.574) | 0.510 (0.485, 0.535) | 0.434 |
| **M2** (Within-market behaviour) | 0.462 (0.441, 0.484) | 0.035 (0.028, 0.044) | 0.962 (0.953, 0.970) | 0.520 (0.499, 0.541) | 0.460 (0.439, 0.482) | 0.593 (0.568, 0.618) | **$1.44x10^{-13}$** |
| **All** (Combined **M1** + **M2**) | 0.546 (0.525, 0.568) | 0.432 (0.411, 0.453) | 0.679 (0.658, 0.699) | 0.611 (0.590, 0.632) | 0.506 (0.484, 0.527) | 0.593 (0.569, 0.618) | **$1.42x10^{-13}$** |
| **Restricted Analysis** | | | | | | | |
| **Almanis-All** | 0.774 (0.764, 0.783) | 0.862 (0.854, 0.869) | 0.674 (0.663, 0.684) | 0.750 (0.740, 0.760) | 0.811 (0.802, 0.820) | 0.838 (0.829, 0.847) | **$<2x10^{-16}$** |
| **NGS2-All** | 0.601 (0.580, 0.622) | 0.709 (0.689, 0.728) | 0.475 (0.453, 0.496) | 0.612 (0.591, 0.633) | 0.583 (0.561, 0.604) | 0.618 (0.594, 0.643) | **$<2x10^{-16}$** |

**Fig. S2. i. Area Under the Curve before and then after trade, stratified by grade of machine-rated quality. ii. Temporal Area Under the Curve gain (95%CI) stratified by grade of machine-rated quality.**

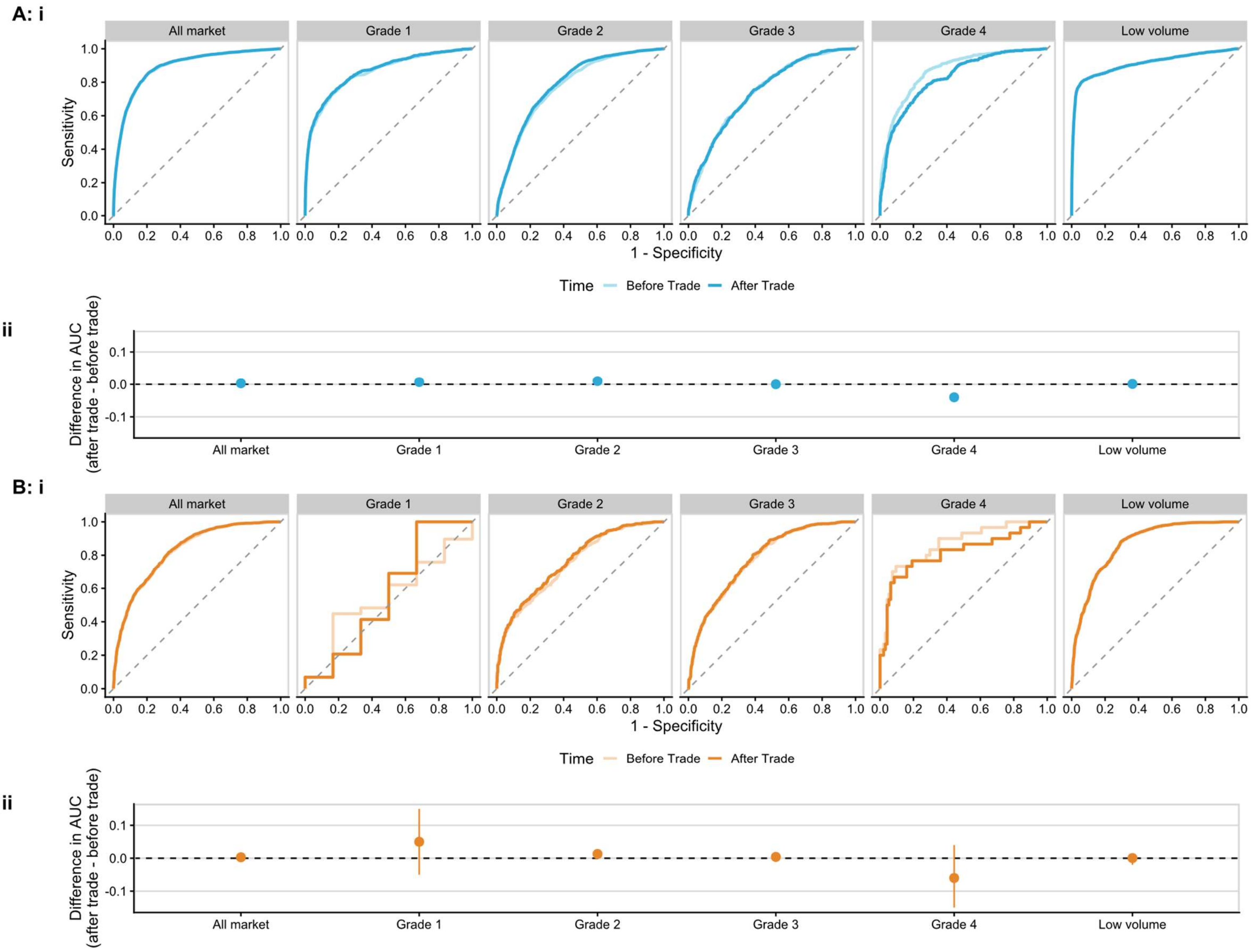

**Panel A: Almanis A. i. Before trade vs after trade AUC (p-value of difference, clustered in market):** All market: *before 0.888 vs after 0.891, p-value=9.13x10*$^{-19}$. Grade 1: *before 0.857 vs after 0.865, p-value=0.001*. Grade 2: *before 0.777 vs after 0.789 p-value=3.64x10*$^{-7}$. Grade 3: before 0.744 vs after 0.746 p-value=0.77. Grade 4*: before 0.868 vs after 0.829 p-value=6.15x10*$^{-9}$. Low volume: before 0.910 vs after 0.910, p-value=0.794. **ii. % AUC Gain after – before trade (95%CI):** All market: *0.03 (0.020, 0.030)* Grade 1: *0.070 (0.03, 1.00)* Grade 2: *1.30 (0.080, 1.80)* Grade 3: 0.20 (-0.80, 1.10) Grade 4: *-3.90 (-5.30, -2.60)*. Low volume: 0.0001 (-9 x 10-4, 0.001).

**Panel B: NGS2. i. Before trade vs after trade AUC, p-value of difference (clustered in market):** All trades*: before 0.824 vs after 0.827, p-value=7.47x10*$^{-6}$. Grade 1: before 0.583 vs after 0.634, p-value=0.33. Grade 2: *before 0.749 vs after 0.761, p-value=0.003*. Grade 3: before 0.781 vs after 0.785 p-value=0.17. Grade 4: before 0.879 vs after 0.822, p-value=0.23. Low volume: before 0.865 vs after 0.866, p-value=0.605. **ii. % AUC Gain after-before trade (95%CI):** All market: *0.003 (0.002, 0.005)* Grade 1: 0.05 (-0.05, 0.15). Grade 2: *0.013 (0.004, 0.021).* Grade 3: 0.004 (-0.002, 0.01). Grade 4: -0.06 (-0.15, 0.04). Low volume: 0.0005 (-0.002, 0.003). **Definitions:** All market: unstratified. Grade 1, 2, 3 and 4 with machine quality ratings of: 0.75 > 1.00, 0.5 > 0.75, 0.25 > 0.5 and 0 > 0.25 respectively. Low volume traders were those whose forecasts did not move the market very much. The cut-off for this classification was a potential maximum relative Brier score of 0.024 on a question, which was equivalent to moving the market forecast by 1.2% when the market probability was 50%.

**Fig. S3. Forecasters vary in the proportion of markets where they demonstrate at least one high quality trade.**

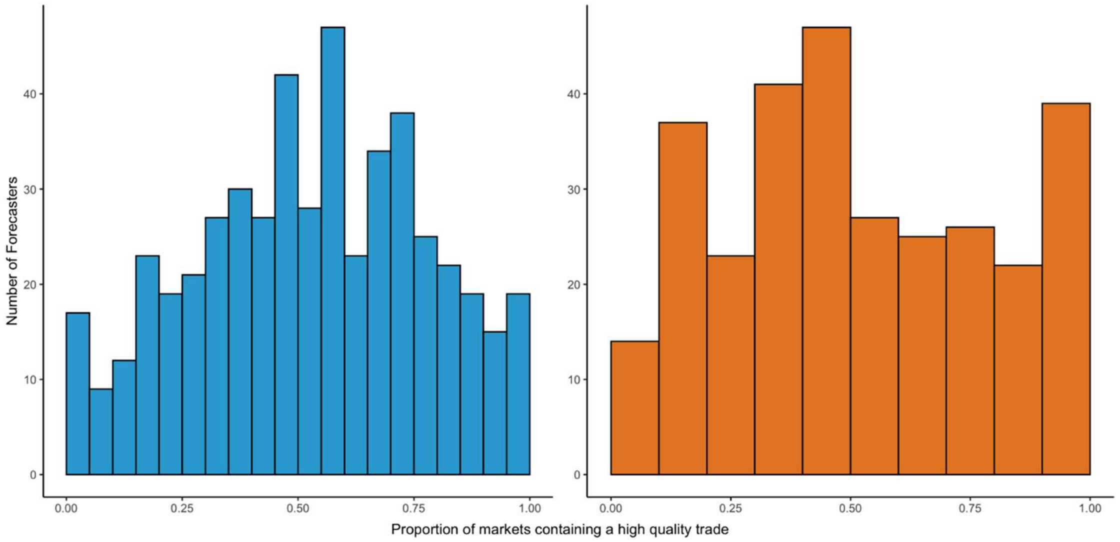

**Note:** The distributions are for forecasters who traded in 5 or more markets. Most forecasters were accurate about 30% to 70% of the time and few very consistently had high quality trades across 90% of the markets they were active in. The median (interquartile range) proportion of a forecaster's markets where a high-quality trade was detected was 0.54 (0.34, 0.72) and 0.50 (0.32,0.75) for Almanis A (blue) and NGS2 (orange) respectively.
